# Comparative Performance of Polygenic Risk Scores for Atherosclerotic Cardiovascular Disease Subtypes in the *All of Us* Research Program

**DOI:** 10.1101/2024.11.05.24316750

**Authors:** Johanna L. Smith, Kristjan Norland, Marwan E. Hamed, Yue Yu, Jie Na, Ozan Dikilitas, Daniel J. Schaid, Iftikhar J. Kullo

**Affiliations:** Department of Cardiovascular Medicine, Mayo Clinic, Rochester, MN, USA; Department of Quantitative Health Sciences, Rochester, MN, USA; Gonda Vascular Center, Mayo Clinic, Rochester, MN, USA

## Abstract

**Background:** Performance and portability of contemporary polygenic risk scores (PRS) for atherosclerotic cardiovascular disease (ASCVD) phenotypes vary based on different methods, training data, and trait ascertainment.

**Objectives:** We aimed to investigate performance and portability of contemporary PRS for ASCVD subtypes: coronary heart disease (CHD), abdominal aortic aneurysm (AAA), ischemic stroke (IS), and peripheral artery disease (PAD), using the *All of Us* Workbench which provides access to a large diverse cohort with phenotype and whole genome sequence data. We also developed and evaluated a multi-trait PRS for each subtype.

**Methods:** Performance of PRS for 4 ASCVD traits and related risk factors was compared across genetic ancestry groups in 245,388 *All of Us* participants. Genetic EUR, African (AFR), Admixed American (AMR), and remaining ancestry groups (combined as Other, OTH) were defined by *All of Us* based on principal components. PRS for CHD, IS, AAA, PAD, and multi-trait (combining PRS for the 4 traits as well as PRS for ASCVD risk factors) were assessed for portability across genetic ancestry groups using hazard ratios (HR) per SD increase.

**Results:** For CHD, CHD_PGS003725_ performed the best (HR for 1 SD increase [95% CI]), across 4 genetic ancestry groups (EUR: 1.72[1.67-1.78], AFR: 1.24[1.18-1.31], AMR: 1.48[1.37-1.59], OTH: 1.65[1.52-1.79]). The best performing PRS for AAA was AAA_PGS003972_ (EUR: 1.68[1.59-1.78], AFR: 1.29[1.13-1.48], AMR: 1.30[1.06-1.60], OTH: 1.45[1.20-1.75]). The best performing IS PRS was IS_PGS000039_ in AFR (1.12[1.06-1.17]), AMR (1.11[1.04-1.19]), and OTH (1.23[1.09-1.38]), and IS_PGS004939_ in EUR (1.16[1.12-1.20]). For PAD, PAD_PGS004940_ performed best in EUR (1.26[1.22-1.30]), AFR (1.11[1.05-1.18]), AMR (1.08[1.01-1.16]), and OTH (1.13[1.04-1.22]). Multi-trait PRS performed better than individual trait PRS for each ASCVD phenotype. Also, PRS derived from multi-ancestry cohorts performed better than those derived from single ancestry.

**Conclusions:** PRS for ASCVD developed from multi-ancestry cohorts and multiple related traits performed best across ancestrally diverse and admixed individuals. PRS for CHD and AAA performed better than those for IS and PAD.

## Background

Atherosclerotic cardiovascular disease (ASCVD) is the leading cause of mortality globally^1,2^ and includes four subtypes: coronary heart disease (CHD), abdominal aortic aneurysm (AAA), ischemic stroke (IS), and peripheral artery disease (PAD)^3^. PRS for predicting risk of ASCVD subtypes^3,4^ are derived from genome-wide association studies (GWAS) of primarily European genetic ancestry (EUR) individuals. Performance of PRS depend on genetic architecture of phenotypes, size, heterogeneity of development and training cohorts, as well as quality of summary statistics^5,6^. Newer PRS aim to include larger GWAS datasets and improve cross-ancestry prediction, resulting in a need to benchmark methods to compare performance in independent and diverse cohorts. In addition, recent studies suggest that linearly combining PRS for multiple related traits as well as training on multi-ancestry data improves prediction^7^.

Herein, we validate and benchmark available PRS for ASCVD phenotypes (i.e., PRS for CHD (PRS_CHD_), PRS for AAA (PRS_AAA_), PRS for IS (PRS_IS_), and PRS for PAD (PRS_PAD_)) in major genetic ancestry populations and explore the utility of newly developed multi-trait and multi-ancestry PRS for each subtype by leveraging data available in the AoU cohort. The *All of Us* (AoU) Researcher Workbench includes short read whole genome sequence data for 245,388 individuals^8^ and corresponding electronic health record (EHR) data^9^. In addition to information regarding ASCVD phenotypes, risk factor data is available for individuals genetically defined as EUR, Middle Eastern (MID), African (AFR), Admixed American (AMR), East Asian (EAS), and South Asian (SAS) ancestry based on principal component information.

We hypothesized that multi-trait PRS would perform better for all ASCVD subtypes given shared genetic architecture of the phenotypes. We evaluated PRS performance portability across genetic ancestry groups^10^.

## Methods

### Data Availability

The general workflow of the study is displayed in Figure 1. Additional information is provided in Supplemental Files (Tables S1-4, Figures S1-7)^3,7,8,11–21^. This study was approved by Mayo Clinic IRB application 10-00278. PRS chosen for validation were publicly available through the PGS Catalog (https://www.pgscatalog.org/), except for two internally developed PRS which will be published on PGS Catalog upon publication (i.e., PGS004939, PGS004940). We used data from the Researcher Workbench for *All of Us* Research Programs Controlled Tier Dataset version 7 and requests for access to data should be made directly with the *All of Us* Research Program.

**Figure 1:**
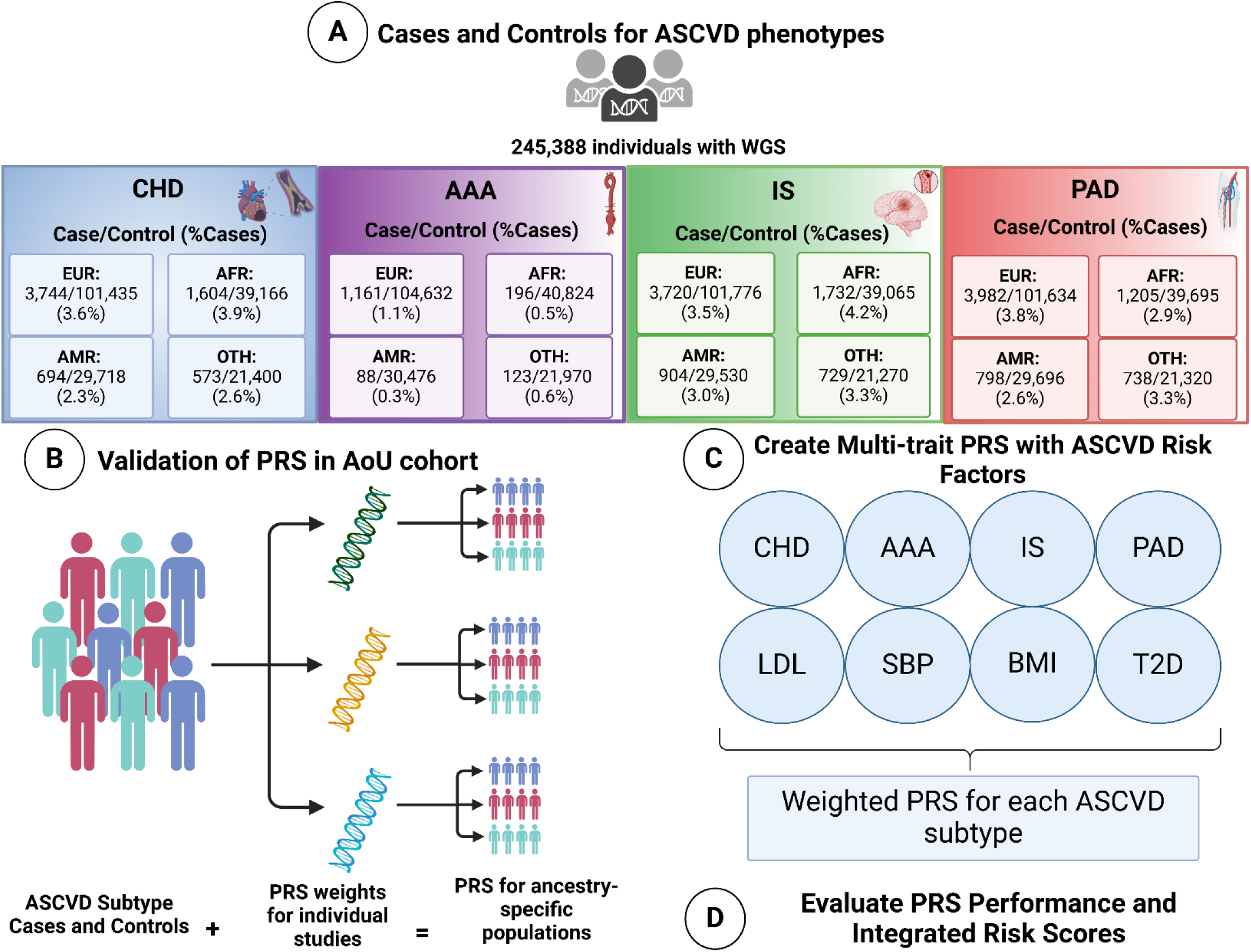
Polygenic risk score (PRS) validation workflow. Demonstrates the creation of cohorts for each ASCVD phenotype, validation of each chosen PRS with the AoU disease-specific cohorts, and statistical analyses that were used to quantify performance of each PRS across all phenotypes.

### Genotype, Phenotype, and Genetic Ancestry Ascertainment

The *All of Us* Researcher Workbench (v7) genomic data released in March 2023 include 245,388 individuals ^8^. We utilized EHR data to ascertain cases and controls for ASCVD subtypes as well as demographic and conventional risk factors. Incident cases for ASCVD subtypes were defined as individuals diagnosed after 6 months since entry into the EHR using algorithms published on pheKB and validated by the eMERGE Network^11^ (https://phekb.org/network-associations/emerge). Controls for ASCVD phenotypes were defined as having no ASCVD diagnosis to date (Table 1).

**Table 1.** ASCVD Phenotype definitions for coronary heart disease (CHD), peripheral artery disease (PAD), abdominal aortic aneurysm (AAA), cerebral vascular disease (CVD), and all ASCVD phenotypes combined.

| ASCVD Phenotype | Case Requirements | Control Requirements |
| --- | --- | --- |
| CHD<sup>*</sup> | 3 separate occurrences of hard diagnostic codes or 1 procedural code | No ASCVD diagnoses |
| AAA<sup>†</sup> | 1 occurrence of a diagnostic or procedural code | No ASCVD diagnoses |
| IS<sup>‡</sup> | 1 occurrence of a diagnostic code | No ASCVD diagnoses |
| PAD<sup>§</sup> | Exclusion of patients with 2 “diagnostic exclusion” codes, then 1 occurrence of a diagnostic or procedural code | No ASCVD diagnoses |

\*CHD- Coronary heart disease
†AAA- Abdominal aortic aneurysm
‡IS- Ischemic stroke
§PAD- Peripheral artery disease

Analyses were restricted to adults (18 years of age or older at first EHR record) and individuals with differing sex at birth and gender designations were excluded (Figure S1). Type 2 Diabetes (T2D), systolic blood pressure (SBP), low-density lipoprotein cholesterol (LDL-C), antihypertensives, and statin use were determined at the most recent data entry before the median time to disease diagnosis for each ancestry group (Table S1) in the control group, or at the time of disease diagnosis for cases^12^.

Genetic ancestry was predetermined by AoU based on principal component analysis and random forest classification using the 1000 Genomes and GNOMAD data. Groups including AFR, AMR, EAS, EUR, MID, SAS, and Other (OTH) were defined in the “ancestry_preds_other” file. We analyzed genetically classified AFR, AMR, and EUR individuals, with the remaining individuals classified as OTH (i.e., highly admixed individuals or those belonging to genetic ancestry groups not large enough to analyze independently)^8^. Hereafter, any reference to ancestry refers to the genetic ancestry derived as stated.

### PRS Performance in AoU

PRS for each subtype were downloaded from the PGS Catalog (www.pgscatalog.org, November 2023), prioritizing recent PRS trained on multi-ancestry populations when available. We included internally developed scores (IS_PGS004939_ and PAD_PGS004940_) due to limited availability for IS and PAD PRS in the PGS Catalog. PRS evaluated in this study are referred to as TRAIT_PGSCatalogReference_ (i.e., PRS_CHD_ by Patel et al. ^13^ is CHD_PGS003725_). Four PRS_CHD_ were identified for testing as more multi-ancestry studies have been published with this phenotype. Two PRS_CHD_ were chosen from one publication^14^ as one of the PRS being implemented in the eMERGE clinical trial^15^. Methods of PRS development with genetic ancestry distributions are outlined in Table 1. PRS for ASCVD risk factors (i.e., T2D, SBP, BMI, LDL-C) were also tested in the AoU cohort. One PRS for each risk factor, (chosen based on recently reported performance and inclusion of large, multi-ancestry cohorts) were used in multi-trait PRS development.

We used the “snp_match” function in the bigsnpR package to match and flip alleles as needed^16^. The “score” function in plink 2 was then used to sum the PRS over all individuals^17^. Testing was performed within genetic ancestry groups (EUR, AFR, AMR) and OTH. We normalized all PGS to zero-mean and unit-variance within each genetic ancestry group.

We followed reporting guidelines^3,18^. We assessed the associations of PRS with ASCVD phenotypes by fitting logistic regression models within each genetic ancestry group, adjusting for age and sex. We tested four different regression models that adjusted for i) age and sex, ii) age, sex, and PRS, iii) Cox proportional hazards using average time to event from first EHR record, sex and PRS, and iv) Cox proportional hazards using average time to event from first EHR record, sex, PRS, and risk factors in each genetic ancestry group (EUR, AFR, AMR, and OTH) for CHD, AAA, IS, PAD, and all ASCVD (Table 3). We additionally estimated OR for subtypes in the top 5% of PRS distribution compared to the rest (Table S2). C-statistics from cox models were reported in Table 3.

C-Statistics (*validate* package) were assessed for a model that included age and sex, as well as a model including age, sex, and PRS (Table S2). PRS results were examined using calibrate and validate functions in the rms package using the bootstrap method with 40 repetitions (Figure S5-8). Nagelkerke and calibration *R*^2^ values were obtained to assess performance of the PRS across genetic ancestry populations and Brier Scores calculated as a measure of accuracy of the predictions (Table S2). Pearson correlations between PRS within each ASCVD phenotype were assessed (Figure S2). HR per SD adjusted for time interval (age at EHR entry to age of diagnosis) and sex were assessed using Cox proportional hazards regression. Schoenfeld residuals were examined for deviations from the proportional-hazards assumption. Adjustments for conventional ASCVD risk factors (i.e., T2D, SBP, LDL-C), as well as statin and antihypertensive use were included in the cox model fit for comparison. Summary information for each PRS validated is in Table 3 and Table S2.

### Multi-trait PRS

We gathered association weights from PRS for T2D, SBP, BMI, and LDL-C in the AoU cohort and included these PRS in an adjusted weight (multi-trait PRS) following previously described methods (Norland et al. 2022)^7^. This method involves linearly combining the trait of interest PRS with risk factor PRS^7^. We utilized one PRS for each ASCVD subtype based on performance (HR per SD): CHD_PGS003356_, AAA_PGS003972_, IS_PGS000039_, and PAD_PGS004940_, excluding PRS that had previously accounted for risk factors included in the analysis (i.e., CHD_PGS003725_ ^13^) to minimize statistical bias. Coefficients to weight the multi-trait PRS components were determined using 10-fold cross validation with *glmnet* and *glmnetUtils* R packages^19^ for each ASCVD phenotype and ancestry group (Figure S#). Multivariable linear regression was performed as previously described for all multi-trait PRS subtypes including the combined ASCVD phenotype.

### Integrated Risk Scores

We also evaluated 10-year ASCVD risk using pooled cohort equations (PCEs)^20^. We then combined PCEs with all PRS to create an integrated risk score (IRS) for each trait to examine accuracy with the equation as follows:

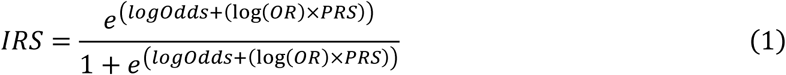

Net reclassification indices (NRI) were evaluated for all PRS and all phenotypes using the *nricens* package with function “nribin()” for categorical and continuous NRI. Continuous and categorical NRI were used to evaluate risk prediction accuracy at a threshold of 10% individuals. Sensitivity and specificity were calculated for all clinical and integrated risk assessments (Supplemental File 2). Risk category reclassifications were also examined based on a threshold of 10-year ASCVD risk of 10% (Figure S9-12).

## Results

Overall, PRS_CHD_ performed best out of all ASCVD subtypes (Figure 2). CHD_PGS003725_ had the strongest associations with CHD in all population-based subsets of the AoU cohort (HR [95% CI]) with the highest performance in EUR (1.72[1.67-1.78]), followed by OTH (1.65[1.52-1.79]), AMR (1.48[1.37-1.59]) and AFR (1.24[1.18-1.31]) (Figure 2, Table S2). The CHD_Multi_ PRS showed similar results to CHD_PGS003725_, with improvements in AFR (1.26[1.20-1.32]) and AMR (1.48[1.37-1.59]) populations, with a decrease in EUR (1.68[1.62-1.73]) and OTH (1.52[1.40-1.65]; Figure 2, Table 2, Table S2). Performance of PRS_CHD_ for AFR populations was much lower (Figure 2, Table 2). Examining the Nagelkerke *R*^2^ values for these validations suggest that the model used to develop CHD_PGS003725_ that integrates other related traits provides better predictive accuracy than CHD_Multi_ (Figure 3, Table S2).

**Figure 2.**
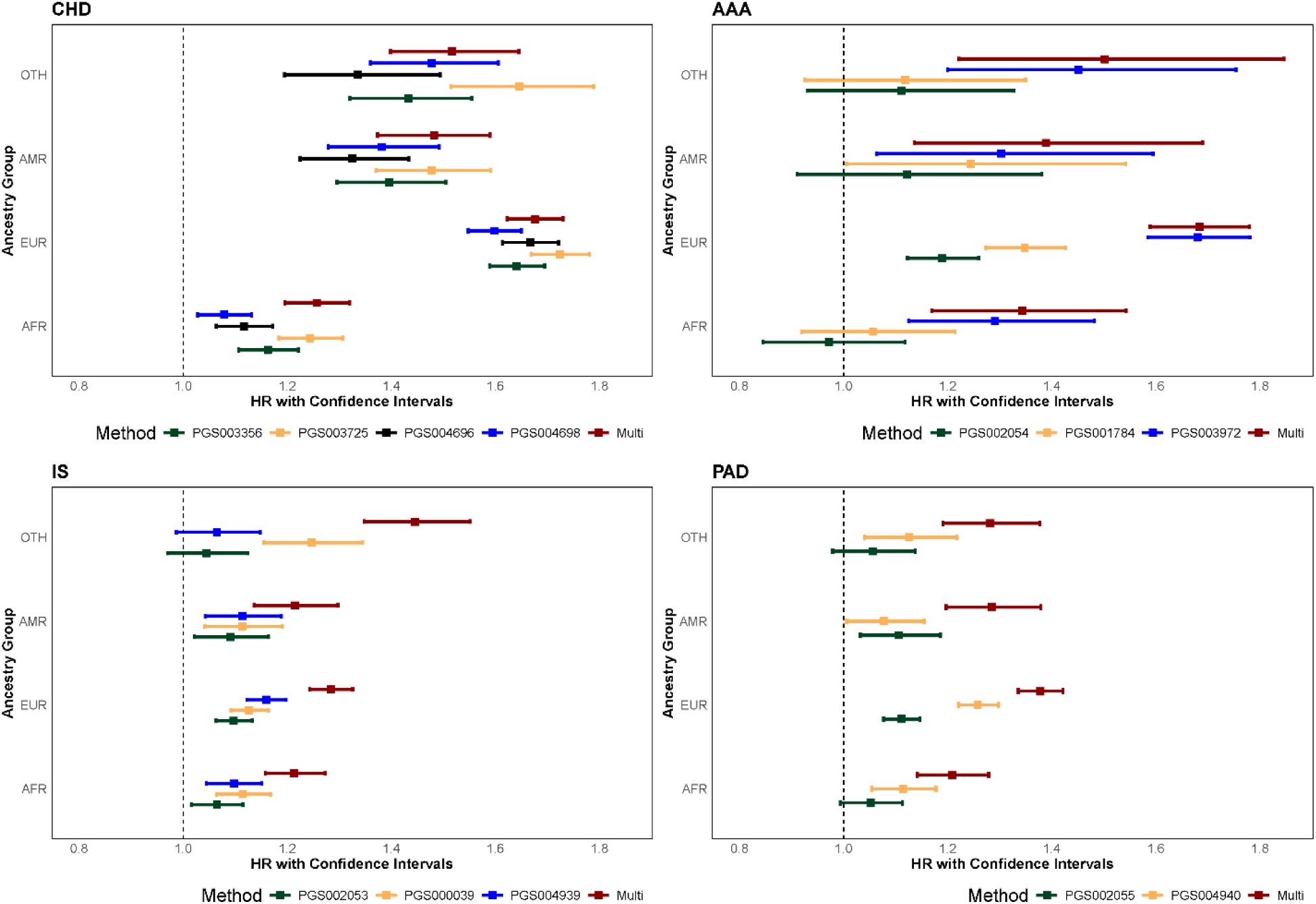
Polygenic risk score validation in African (AFR), European (EUR), Admixed American (AMR), and Other (OTH) genetic ancestry populations for coronary heart disease (CHD), cerebrovascular disease (CVD), abdominal aortic aneurysm (AAA), and peripheral artery disease (PAD). Plots show odds ratio per standard deviation (OR per SD) with confidence intervals for each PRS tested in each phenotype.

**Figure 3.**
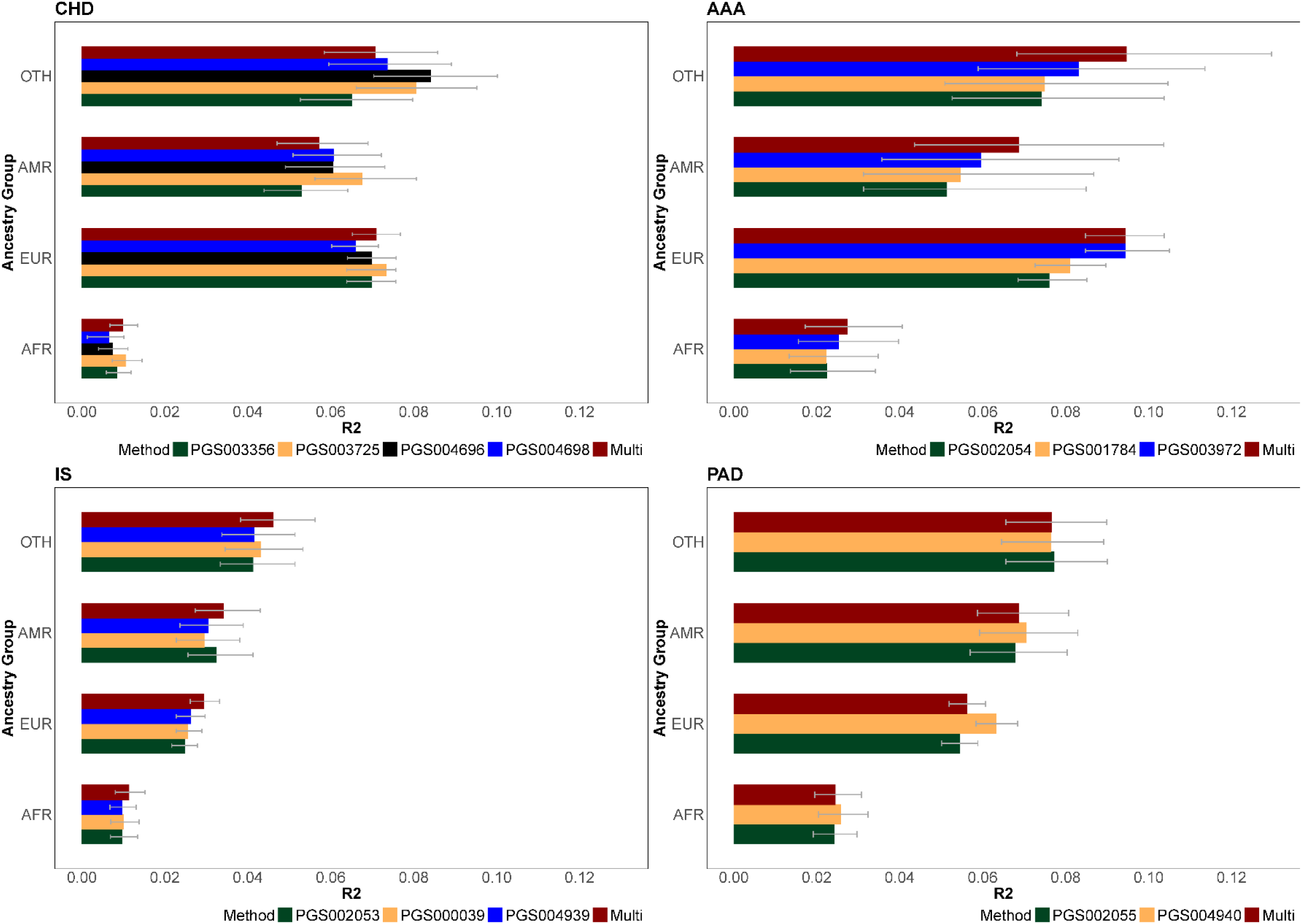
Nagelkerke R^2^ values with 95% CI of PRS for CHD, IS, AAA, and PAD for single trait PRS and multi-trait PRS for African (AFR), European (EUR), Admixed American (AMR), and Other (OTH) genetic ancestry groups.

**Table 2.**
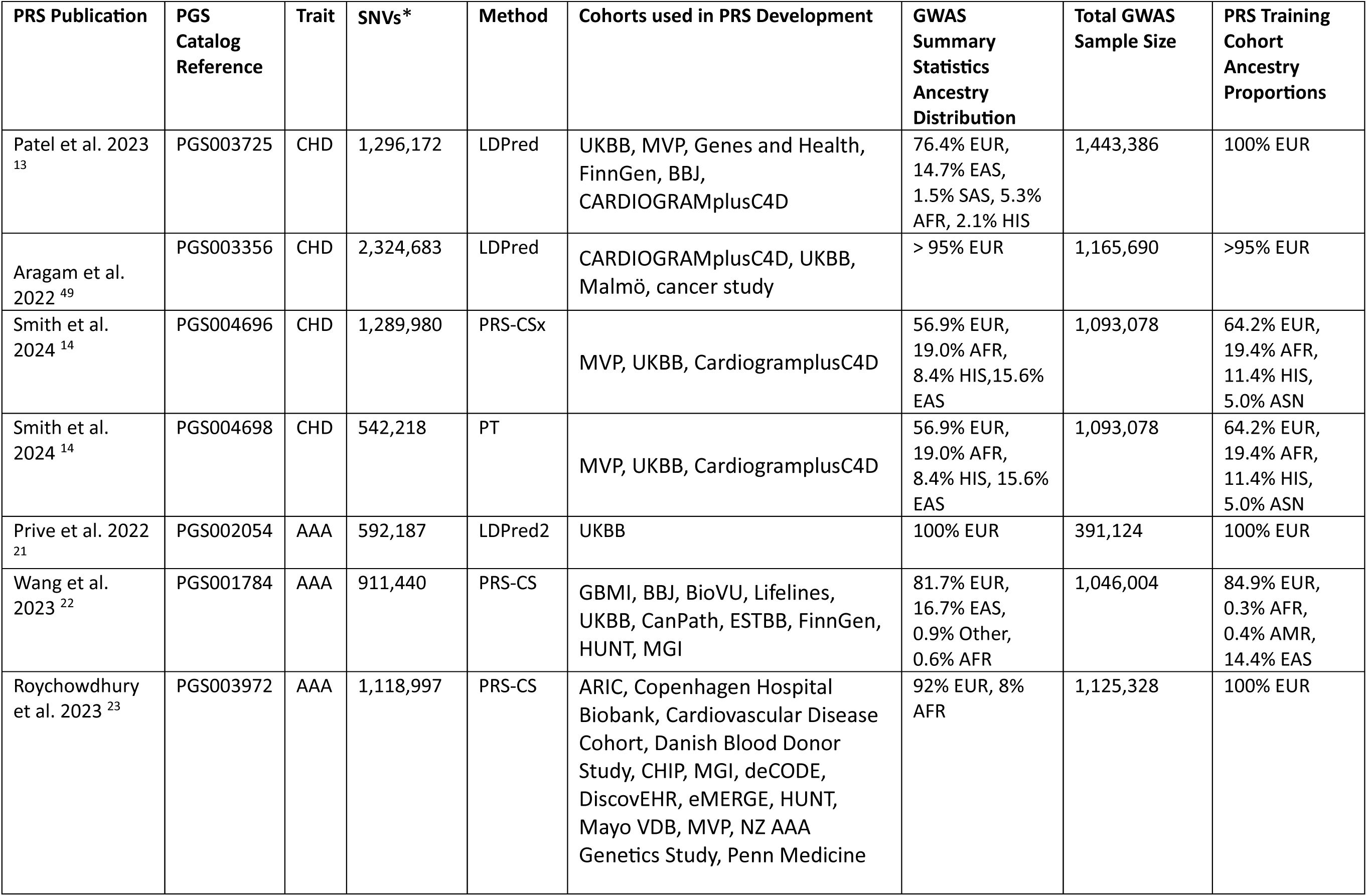
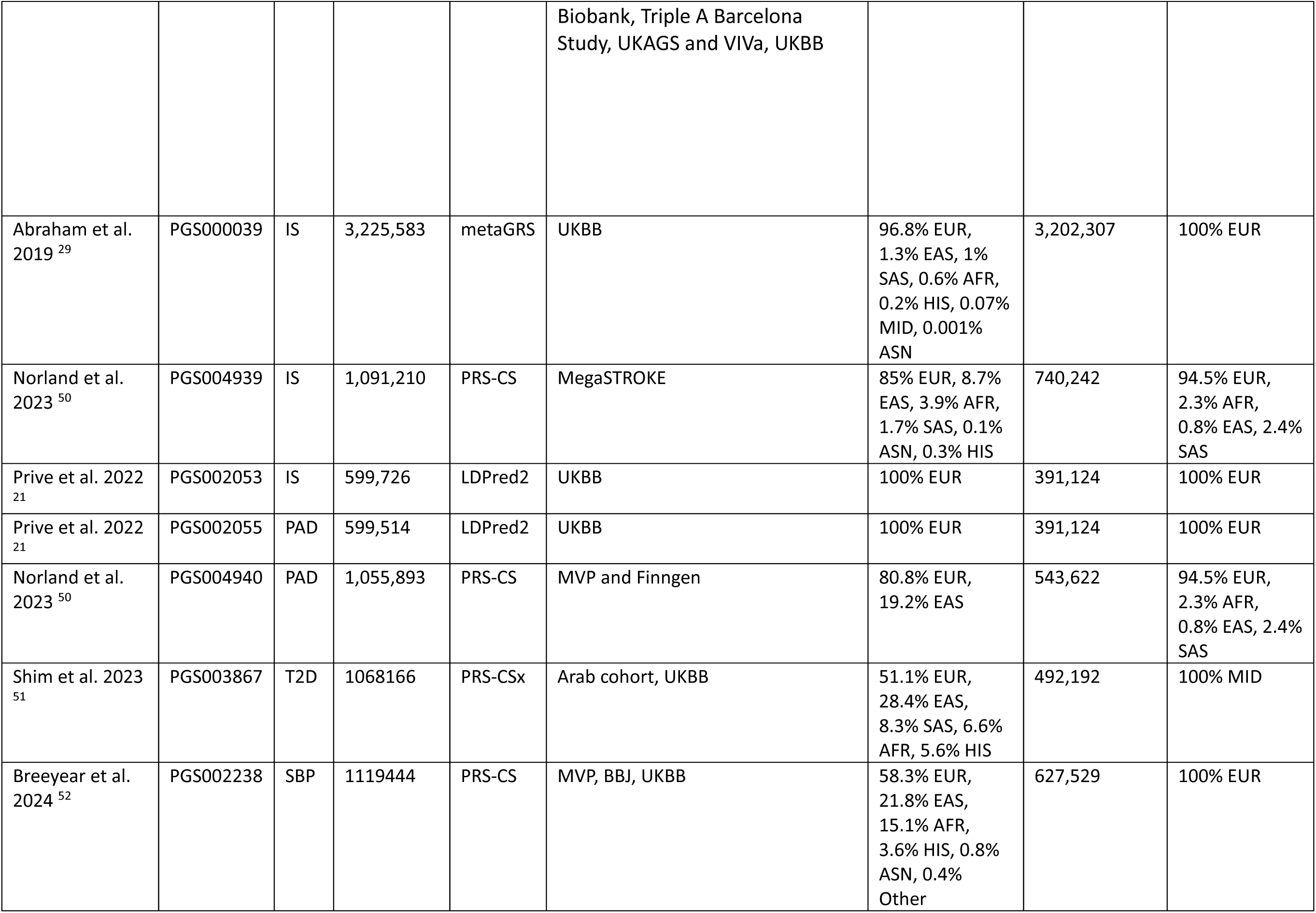
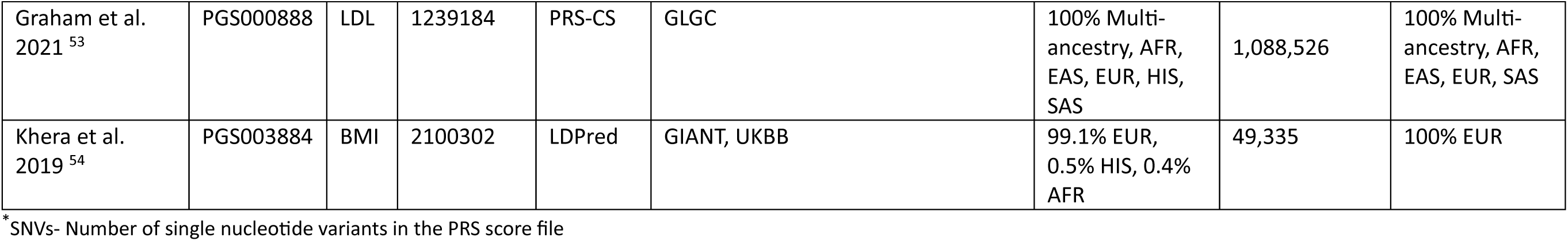
Polygenic risk scores from PGS Catalog that were tested in the All of Us cohort with general PRS development method, number of single nucleotide variants (SNVs) included, and training cohorts listed for each PRS noting if multi-ancestry cohorts were used in PRS development.

**Table 3.** Performance of PRS for ASCVD phenotypes including hazard ratios (HR per SD) (adjusted for age and sex) and HR adjusted for age, sex, and contemporary risk factors (diabetes, systolic blood pressure, LDL-C, antihypertensive medication, and statin medication) for each genetic ancestry group and all ancestries combined. Base model refers to adjustments for age and sex variables.

| PRS | Ancestry | HR per SD (95% CI)<br>base + PRS<sup>*</sup> | P-Value | C-Statistic<br>base<sup>†</sup> | C-Statistic<br>base + PRS<sup>‡</sup> | HR per SD (95% CI)<br>base + PRS + Risk Factors<sup>§</sup> | P-Value | C-Statistic<br>base + PRS +<br>Risk Factors |
| --- | --- | --- | --- | --- | --- | --- | --- | --- |
| Coronary Heart Disease |  |  |  |  |  |  |  |  |
| CHD<sub>PGS003356</sub> | EUR | 1.641 (1.589-1.695) | 5.88E-201 | 0.729 | 0.763 | 1.553 (1.503-1.604) | 6.48E-156 | 0.793 |
|  | AFR | 1.163 (1.107-1.222) | 1.58E-09 | 0.651 | 0.655 | 1.130 (1.075-1.187) | 1.28E-06 | 0.718 |
|  | AMR | 1.396 (1.296-1.505) | 1.67E-18 | 0.755 | 0.769 | 1.300 (1.207-1.401) | 5.19E-12 | 0.808 |
|  | OTH | 1.433 (1.321-1.555) | 6.20E-18 | 0.768 | 0.781 | 1.365 (1.257-1.482) | 1.48E-13 | 0.797 |
| CHD<sub>PGS003725</sub> | EUR | 1.724 (1.669-1.781) | 1.24E-236 | 0.729 | 0.771 | 1.584 (1.532-1.637) | 8.47E-164 | 0.796 |
|  | AFR | 1.244 (1.184-1.307) | 4.34E-18 | 0.651 | 0.661 | 1.166 (1.109-1.225) | 1.43E-09 | 0.719 |
|  | AMR | 1.477 (1.371-1.591) | 1.08E-24 | 0.755 | 0.770 | 1.395 (1.295-1.503) | 2.37E-18 | 0.809 |
|  | OTH | 1.646 (1.515-1.789) | 4.57E-32 | 0.768 | 0.800 | 1.527 (1.404-1.660) | 4.04E-23 | 0.811 |
| CHD<sub>PGS004696</sub> | EUR | 1.667 (1.614-1.722) | 5.57E-212 | 0.729 | 0.766 | 1.552 (1.502-1.603) | 2.39E-154 | 0.795 |
|  | AFR | 1.117 (1.064-1.172) | 8.44E-06 | 0.651 | 0.654 | 1.082 (1.030-1.135) | 1.52E-03 | 0.716 |
|  | AMR | 1.325 (1.225-1.434) | 2.52E-12 | 0.755 | 0.761 | 1.381 (1.278-1.493) | 3.60E-16 | 0.808 |
|  | OTH | 1.336 (1.195-1.494) | 3.53E-07 | 0.768 | 0.774 | 1.344 (1.203-1.501) | 1.55E-07 | 0.796 |
| CHD<sub>PGS004698</sub> | EUR | 1.598 (1.548-1.650) | 1.19E-179 | 0.729 | 0.762 | 1.499 (1.451-1.548) | 4.93E-131 | 0.791 |
|  | AFR | 1.079 (1.028-1.132) | 2.00E-03 | 0.651 | 0.652 | 1.050 (1.001-1.102) | 4.51E-02 | 0.715 |
|  | AMR | 1.382 (1.279-1.492) | 7.68E-01 | 0.755 | 0.768 | 1.409 (1.305-1.521) | 1.93E-18 | 0.810 |
|  | OTH | 1.478 (1.360-1.606) | 3.41E-20 | 0.768 | 0.784 | 1.399 (1.287-1.520) | 3.31E-15 | 0.799 |
| CHD<sub>Multi</sub> | EUR | 1.676 (1.623-1.731) | 1.30E-218 | 0.729 | 0.768 | 1.545 (1.496-1.597) | 5.34E-151 | 0.794 |
|  | AFR | 1.257 (1.196-1.322) | 1.97E-19 | 0.651 | 0.662 | 1.168 (1.110-1.229) | 1.80E-09 | 0.719 |
|  | AMR | 1.482 (1.374-1.598) | 1.75E-24 | 0.755 | 0.773 | 1.272 (1.178-1.373) | 8.28E-10 | 0.806 |
|  | OTH | 1.517 (1.399-1.645) | 4.37E-24 | 0.768 | 0.789 | 1.422 (1.310-1.543) | 4.00E-17 | 0.802 |
| Abdominal Aortic Aneurysm |  |  |  |  |  |  |  |  |
| AAA<sub>PGS002054</sub> | EUR | 1.190 (1.123-1.260) | 3.20E-09 | 0.824 | 0.826 | 1.181 (1.115-1.251) | 1.32E-08 | 0.833 |
|  | AFR | 0.972 (0.845-1.118) | 6.92E-01 | 0.768 | 0.769 | 0.965 (0.839-1.109) | 6.12E-01 | 0.809 |
|  | AMR | 1.122 (0.911-1.381) | 0.278 | 0.820 | 0.818 | 1.108 (0.897-1.369) | 3.42E-01 | 0.841 |
|  | OTH | 1.111 (0.931-1.327) | 0.244 | 0.824 | 0.826 | 1.112 (0.931-1.329) | 2.43E-01 | 0.825 |
| AAA<sub>PGS001784</sub> | EUR | 1.348 (1.273-1.427) | 1.51E-24 | 0.824 | 0.830 | 1.336 (1.262-1.415) | 4.48E-23 | 0.837 |
|  | AFR | 1.057 (0.920-1.214) | 0.436 | 0.768 | 0.769 | 1.029 (0.896-1.181) | 6.89E-01 | 0.809 |
|  | AMR | 1.245 (1.006-1.542) | 0.044 | 0.820 | 0.820 | 1.188 (0.960-1.469) | 1.13E-01 | 0.841 |
|  | OTH | 1.118 (0.925-1.350) | 0.248 | 0.824 | 0.823 | 1.098 (0.908-1.327) | 3.35E-01 | 0.822 |
| AAA<sub>PGS003972</sub> | EUR | 1.680 (1.585-1.781) | 3.55E-68 | 0.824 | 0.842 | 1.636 (1.544-1.734) | 1.48E-61 | 0.848 |
|  | AFR | 1.291 (1.125-1.482) | 2.80E-04 | 0.768 | 0.768 | 1.261 (1.096-1.451) | 1.20E-03 | 0.807 |
|  | AMR | 1.303 (1.064-1.595) | 1.00E-02 | 0.820 | 0.818 | 1.254 (1.028-1.530) | 2.58E-02 | 0.842 |
|  | OTH | 1.451 (1.200-1.754) | 1.24E-04 | 0.824 | 0.840 | 1.438 (1.189-1.739) | 1.77E-04 | 0.839 |
| AAA<sub>Multi</sub> | EUR | 1.684 (1.589-1.783) | 9.99E-69 | 0.824 | 0.842 | 1.637 (1.544-1.735) | 1.47E-61 | 0.848 |
|  | AFR | 1.344 (1.170-1.543) | 2.81E-05 | 0.768 | 0.769 | 1.320 (1.146-1.521) | 2.21E-08 | 0.808 |
|  | AMR | 1.389 (1.136-1.699) | 1.37E-03 | 0.820 | 0.825 | 1.431 (1.169-1.752) | 5.15E-04 | 0.848 |
|  | OTH | 1.506 (1.222-1.846) | 1.14E-04 | 0.824 | 0.837 | 1.548 (1.256-1.908) | 4.21E-05 | 0.838 |
| Ischemic Stroke |  |  |  |  |  |  |  |  |
| IS<sub>PGS002053</sub> | EUR | 1.097 (1.063-1.133) | 1.26E-08 | 0.722 | 0.724 | 1.079 (1.045-1.114) | 3.34E-06 | 0.762 |
|  | AFR | 1.065 (1.016-1.115) | 8.44E-03 | 0.682 | 0.683 | 1.042 (0.994-1.092) | 8.68E-02 | 0.763 |
|  | AMR | 1.091 (1.022-1.164) | 8.89E-03 | 0.731 | 0.730 | 1.073 (1.005-1.145) | 3.38E-02 | 0.757 |
|  | OTH | 1.045 (0.970-1.125) | 0.245 | 0.719 | 0.719 | 1.019 (0.946-1.097) | 6.22E-01 | 0.745 |
| IS<sub>PGS000039</sub> | EUR | 1.127 (1.092-1.164) | 2.08E-13 | 0.722 | 0.725 | 1.086 (1.052-1.122) | 4.72E-07 | 0.762 |
|  | AFR | 1.115 (1.064-1.168) | 4.99E-06 | 0.682 | 0.685 | 1.071 (1.022-1.123) | 4.34E-03 | 0.763 |
|  | AMR | 1.114 (1.042-1.191) | 1.47E-03 | 0.731 | 0.732 | 1.027 (0.961-1.099) | 4.30E-01 | 0.758 |
|  | OTH | 1.247 (1.156-1.345) | 1.06E-08 | 0.719 | 0.727 | 1.158 (1.072-1.252) | 2.14E-04 | 0.749 |
| IS<sub>PGS004939</sub> | EUR | 1.160 (1.123-1.198) | 1.367E-19 | 0.722 | 0.726 | 1.124 (1.088-1.161) | 1.39E-12 | 0.763 |
|  | AFR | 1.098 (1.048-1.151) | 8.61E-05 | 0.682 | 0.684 | 1.054 (1.005-1.104) | 2.92E-02 | 0.763 |
|  | AMR | 1.114 (1.043-1.189) | 1.23E-03 | 0.731 | 0.732 | 1.037 (0.972-1.107) | 2.73E-01 | 0.758 |
|  | OTH | 1.065 (0.987-1.148) | 0.1067 | 0.719 | 0.719 | 1.044 (0.968-1.125) | 2.65E-01 | 0.745 |
| IS<sub>Multi</sub> | EUR | 1.284 (1.243-1.326) | 6.19E-52 | 0.722 | 0.734 | 1.162 (1.124-1.200) | 5.05E-19 | 0.765 |
|  | AFR | 1.214 (1.158-1.273) | 8.02E-16 | 0.682 | 0.692 | 1.107 (1.056-1.162) | 3.07E-05 | 0.764 |
|  | AMR | 1.215 (1.137-1.297) | 6.86E-09 | 0.731 | 0.736 | 1.062 (0.992-1.136) | 8.21E-02 | 0.759 |
|  | OTH | 1.230 (1.093-1.384) | 5.894E-04 | 0.719 | 0.763 | 1.282 (1.191-1.380) | 3.94E-11 | 0.757 |
| Peripheral Artery Disease |  |  |  |  |  |  |  |  |
| PAD<sub>PGS002055</sub> | EUR | 1.111 (1.077-1.147) | 2.83E-11 | 0.765 | 0.767 | 1.088 (1.055-1.123) | 1.00E-07 | 0.798 |
|  | AFR | 1.052 (0.994-1.113) | 8.03E-02 | 0.748 | 0.748 | 1.031 (0.974-1.091) | 2.95E-01 | 0.801 |
|  | AMR | 1.106 (1.032-1.186) | 4.59E-03 | 0.770 | 0.771 | 1.043 (0.973-1.117) | 2.40E-01 | 0.807 |
|  | OTH | 1.056 (0.980-1.138) | 1.56E-01 | 0.785 | 0.786 | 1.034 (0.959-1.114) | 3.90E-01 | 0.798 |
| PAD<sub>PGS004940</sub> | EUR | 1.258 (1.221-1.298) | 3.10E-49 | 0.765 | 0.774 | 1.269 (1.229-1.309) | 2.96E-49 | 0.804 |
|  | AFR | 1.114 (1.054-1.178) | 1.40E-04 | 0.748 | 0.749 | 1.117 (1.055-1.183) | 1.43E-04 | 0.802 |
|  | AMR | 1.078 (1.006-1.155) | 3.20E-02 | 0.770 | 0.771 | 1.096 (1.020-1.177) | 1.24E-02 | 0.809 |
|  | OTH | 1.126 (1.040-1.218) | 3.28E-03 | 0.785 | 0.786 | 1.185 (1.100-1.276) | 8.03E-06 | 0.802 |
| PAD<sub>Multi</sub> | EUR | 1.378 (1.336-1.422) | 2.63E-90 | 0.765 | 0.780 | 1.159 (1.123-1.195) | 2.32E-20 | 0.800 |
|  | AFR | 1.209 (1.142-1.279) | 5.23E-11 | 0.748 | 0.753 | 1.088 (1.029-1.151) | 3.14E-03 | 0.801 |
|  | AMR | 1.285 (1.197-1.379) | 3.77E-12 | 0.770 | 0.778 | 1.032 (0.964-1.104) | 3.66E-01 | 0.807 |
|  | OTH | 1.281 (1.191-1.377) | 2.38E-11 | 0.785 | 0.793 | 1.046 (0.969-1.129) | 2.48E-01 | 0.798 |

\* HR per SD (95% CI)- Hazard ratio per standard deviation with 95% confidence interval adjusting for the base model of age and sex in addition to PRS
†C-Statistic<sub>base</sub> - Concordance statistic of the base model of age and sex
‡ C-Statistic<sub>base + PRS</sub>- Concordance statistic of the base model of age and sex in addition to PRS
§ HR per SD (95% CI)<sub>base + PRS + Risk Factors</sub> - Hazard ratio per standard deviation with 95% confidence interval adjusting for the base model of age, sex, PRS, and risk factors including diabetes, systolic blood pressure, LDL-C, antihypertensive medication, and statin medication
|| C-Statistic<sub>base + PRS + Risk Factors</sub>- Concordance statistic of the base model of age, sex, PRS, and risk factors including diabetes, systolic blood pressure, LDL-C, antihypertensive medication, and statin medication

We compared three PRS_AAA_ (i.e., AAA_PGS001784_, and AAA_PGS003972_) in the 4 ancestry groups in AoU. AAA_PGS002054_ was developed using LDPred2, and the remaining two PRS were developed with PRS-CS^21–23^. AAA_PGS001784_ and AAA_PGS003972_ were developed using EUR cohorts, and both utilized multi-ancestry GWAS summary statistics (Table 2)^22,23^. PRS-CS scores outperformed the LDPred2 score in all ancestry groups, though it is noteworthy to state GWAS summary statistics used in training data differed (Figure 2-3, Table S2). AAA_PGS003972_ has the largest GWAS training data for AAA and was the best performing PRS for AAA across ancestry groups with HR per SD of 1.68[1.59-1.78] for EUR, 1.45[1.20-1.75] for OTH, 1.29[1.13-1.48] for AFR, and 1.30[1.06-1.60] for AMR, despite limitations in training diversity. AAA_Multi_ increased calibration and Nagelkerke R^2^ value of the single trait PRS by an average of 18.5% (Table S2).

Three PRS ^24,25^ varying in linkage disequilibrium adjustment methods were compared using the AoU cohort^26–30^. The PRS_IS_ were trained on EUR populations, limiting the portability of these scores between ancestry groups, however, IS_PGS000039_ and IS_PGS004939_ utilized multi-ancestry GWAS summary statistics^7,29^, improving the cross-ancestry performance compared to IS_PGS000053_. There was also a difference in PRS development method, allowing comparison of metaGRS as a multi-trait method and PRS-CS as a single-trait continuous shrinkage method (IS_PGS000039_ and IS_PGS004939_)^28,29,31^. For EUR populations, IS_PGS004939_ performed best (1.16[1.12-1.20]), whereas IS_PGS000039_ performed best for OTH (1.25[1.16-1.35]), AFR (1.12[1.06-1.17]), and AMR (1.11[1.04-1.19]; Figure 2-3, Table S2). IS_Multi_ performed better in the 4 ancestry groups (EUR: 1.28[1.24-1.33], AFR: 1.21[1.16-1.27], AMR: 1.22[1.14-1.30], OTH: 1.45[1.35-1.55]) with an average increase of HR per SD 1.16-fold.

Portability of the PRS_PAD_ was limited overall with essentially no predictive value for AFR (HR for 1 SD ∼1 with large standard errors). PAD_PGS004940_ consistently outperformed PAD_PGS002055_ for EUR (1.26[1.22-1.30]), OTH (1.13[1.04-1.22]), and AFR (1.11[1.05-1.18]) and PAD_PGS002055_ performed best for AMR (1.11[1.03-1.19]; Figure 2, Table S2). However, PAD_Multi_ performed better with a HR per SD increase of 9.5% for EUR, 8.5% for AFR, 16% for AMR, and 13.8% for OTH from the best performing PRS.

Adjustment for conventional risk factors [type II diabetes (T2D), systolic blood pressure (SBP), low-density lipoprotein cholesterol (LDL-C)], as well as antihypertensive and statin use^12^ attenuated HR per SD for the PRS (∼0.1 HR per SD decrease), while increasing C-statistic (∼0.1 C-statistic increase). Overall, the multi-trait PRS for each ASCVD phenotype performed better than the single trait PRS, supporting the inclusion of ASCVD subtypes and risk factors in PRS development. However, the multi-trait PRS for each trait was trained and tested using *AoU* and results may be inflated. Analyses for OTH expectedly showed varying performance across ASCVD subtypes and individual PRS for performance statistics as OTH includes a broad variety of admixed and non-admixed individuals with limited sample sizes and resembled the performance for AMR. Calibrations and correlations for each PRS, trait, and ancestry group showed variable results relative to phenotype and ancestry group (Figures S5-8). R^2^ statistics from calibration are shown in Table S2 and Nagelkerke R^2^ with 95% CI are displayed in Figure 3. As EUR had larger training data sets, PRS performed better in EUR than in other ancestries for all ASCVD subtypes. However, EUR-trained PRS continue to perform poorly in AFR^32^.

We also examined performance of integrated risk scores (IRS) that included PRS and the pooled cohort equation (PCE) to estimate 10-year ASCVD risk in the *AoU* cohort. PRS were included as shown in Equation 1 (Methods). C-Statistics for all ASCVD subtype IRS indicate reasonable discrimination (0.682-0.778; Table S7). We identified changes in risk categorization for individuals using a 10% 10-year ASCVD risk threshold as shown in Figures S8-11, as well as continuous and categorical NRI (Table S7). IRS showed improvement of prediction for 10-year ASCVD risk scores. Predictive performance of PRS varied across subtypes. Multi-trait and multi-ancestry PRS had better predictive performance than scores derived from single ancestry.

## Discussion

Performance of contemporary PRS for ASCVD subtypes varied across traits, ancestry groups, and development methods. PRS for CHD and AAA subtypes performed better across genetic ancestry groups than IS and PAD (Figures 2, 3). Expectedly, performance was best in those of EUR ancestry. Multi-ancestry and multi-trait PRS tended to perform better. Lower performance in AFR and admixed populations was consistent across all analyses.

PRS_CHD_ have evolved and improved over the last decade^33^ but remain suboptimal for non-EUR groups. For example, the LDPred2-based method used to develop CHD_PGS003725_^13^ combined GWAS summary statistics for CHD in addition to other ASCVD phenotypes and risk factors, then was trained on primarily EUR populations in the UK Biobank^13^. This method had comparable performance to the multi-trait PRS developed in this study. Expanding the multi-trait method to other ASCVD subtypes increased performance, indicating that leveraging genetic correlations between other traits improves performance for each ASCVD trait compared to single-trait scores^34–37^. Despite the performance gains using multi-trait methods, PRS for non-EUR groups remain suboptimal.

PRS_PAD_ and PRS_IS_ performed less well, implying need for more data and diversity for these phenotypes before implementation into clinical practice (Figure 3). PRS_PAD_ performs similarly for AMR and OTH but has a lower average R^2^ for EUR than PRS_CHD_ and PRS_AAA_ with broader confidence intervals overall (Figure 3). HR per SD of up to 1.69 (IS) and OR per SD of 1.44 (PAD) have been reported for cohort-based studies, however, the scores were not externally tested and were not available publicly or on PGS Catalog^38,39^. As many publications reporting metrics for PRS preceded or did not adhere the current guidelines^3,18^, direct comparison of PRS performance is difficult. Additionally, PAD is a heterogenous phenotype, and case ascertainment may vary across cohorts. Variable phenotyping can lead to variability in heritability estimates as well. For instance, heritability for PAD has been reported from modest (11%)^38^ to moderate (55%)^40^.

Differential performance of PRS for an ASCVD subtype across genetic ancestry groups could be due to varying number of cases across population groups, case ascertainment algorithms within GWAS and PRS development, PRS development methods, and overall diversity of training populations. Consistently lower performances of PRS for AFR is likely due to the greater genetic differences from EUR, which compose most training data available at varying stages of PRS development process^13,14^. Whether differences of causal variants and subsequent effect sizes contribute to performance is unclear^41,42^. As in other traits, PRS for ASCVD perform less well for AFR, highlighting the need for inclusion of more diverse individuals and improving portability of scores through methods to mitigate performance variability^43^.

IRS results (Table S4) had similar trends of performance across genetic ancestry groups and subtype (Table S2). It should be noted that PCEs include self-reported race designations^10^, but risk is likely better estimated by using social determinants of health methods instead of race when they become more available^44^.

### Clinical Implications

The *All of Us* cohort is useful for validation of contemporary PRS as it includes a large, diverse group of individuals and has not yet been included in training datasets, thus avoiding overfitting. PRS_AAA_ and PRS_CHD_ performed robustly, suggesting potential for clinical utility. However, PRS for IS and PAD performed less well. Previous use of PRS for ASCVD phenotypes in clinical settings have shown promising results. For example in the MI-GENES trial, patients choose to implement changes based on PRS to reduce risk of CHD by including statins and/or healthier lifestyles^45^. The Electronic Medical Records and Genomics (eMERGE) Network is testing clinical implementation^15^ of PRS for 9 common conditions^46^. Performance of PRS for ASCVD phenotypes is limited for AFR^10,46^. However, the process of benchmarking in a highly diverse cohort enabled the identification of best-performing PRS for use in clinical risk scores. IRS used for clinical risk evaluation showed an increase in performance, when integrating PRS_CHD_ (Table S4). Therefore, IRS with the best performing PRS will provide the most accurate risk estimates for patients.

### Limitations

The AoU cohort defined genetic ancestry based on projection of samples onto the PC space, rather than more accurate methods of determining genetic ancestry such as ADMIXTURE and SCOPE, which may have labeled some individuals differently^47,48^. Additionally, few individuals present in the AoU cohort could also be subjects in other study cohorts used in development of the PRS weights.

## Conclusions

PRS for ASCVD developed from multi-ancestry cohorts and multiple related traits performed best across ancestrally diverse and admixed individuals. PRS for CHD and AAA performed better than those for IS and PAD. Limited sample sizes and diversity within development cohorts restrict the portability of these PRS. As PRS continue to evolve, periodic assessment of performance is necessary. This study highlights the need for diverse cohorts for GWAS and PRS development for all ASCVD subtypes.

## Data Availability

All data produced in the present work are contained in the manuscript.
Individual-level data is available upon request or application to All of Us Research Program.

## Non-standard Abbreviations and Acronyms

PRS: Polygenic risk score
IRS: Integrated risk score
AoU: *All of Us*
ASCVD: Atherosclerotic cardiovascular disease
CHD: Coronary heart disease
AAA: Abdominal aortic aneurysm
IS: Ischemic stroke
PAD: Peripheral artery disease
EUR: European genetic ancestry
AFR: African genetic ancestry
AMR: Admixed American genetic ancestry
OTH: Other genetic ancestry
GWAS: Genome-wide association study
PRS_CHD_: Polygenic risk score for coronary heart disease
PRS_AAA_: Polygenic risk score for abdominal aortic aneurysm
PRS_IS_: Polygenic risk score for ischemic stroke
PRS_PAD_: Polygenic risk score for peripheral artery disease
Multi: Multiple trait inclusion
eMERGE Network: electronic Medical Records and Genomics Network
PGS Catalog: Polygenic Score Catalog
LDL-C: Low-density lipoprotein cholesterol
SBP: Systolic blood pressure
T2D: Type II diabetes
BMI: Body mass index
OR per SD: Odds ratio per standard deviation increase
HR per SD: Hazard ratio per standard deviation increase
Top 5% OR vs rest: Odds ratio for the top 5% risk individuals versus the rest of the cohort
CI: Confidence interval
C-Statistic: Concordance Statistic
NRI: Net reclassification index
AUC: Area under the operating curve
SNV: Single nucleotide variant

## Acknowledgements

We acknowledge the *All of Us* participants for contributions to the *All of Us* Researcher Workbench for the opportunity to contribute this research. We thank the National Institute of Health’s *All of Us* Research Program for making the data available for use in this study.

## References

1. Mensah GA, Fuster V, Murray CJL, Roth GA, Collaborators GBoCDaR. Global Burden of Cardiovascular Diseases and Risks, 1990-2022. J American Coll Cardiol. 2023;82:2350–2473. doi: 10.1016/j.jacc.2023.11.007

2. Roth GA, Nguyen G, Forouzanfar MH, Mokdad AH, Naghavi M, Murray CJL. Estimates of Global and Regional Premature Cardiovascular Mortality in 2025. Circulation. 2015;132:1270–1282. doi: 10.1161/circulationaha.115.016021

3. Smith JL, Schaid DJ, Kullo IJ. Implementing Reporting Standards for Polygenic Risk Scores for Atherosclerotic Cardiovascular Disease. Current Atherosclerosis Reports. 2023;25:323–330. doi: 10.1007/s11883-023-01104-3

4. Kullo IJ, Lewis CM, Inouye M, Martin AR, Ripatti S, Chatterjee N. Polygenic scores in biomedical research. Nature Reviews Genetics. 2022;23:524–532. doi: 10.1038/s41576-022-00470-z

5. Wang Y, Tsuo K, Kanai M, Neale BM, Martin AR. Challenges and Opportunities for Developing More Generalizable Polygenic Risk Scores. Annual Review of Biomedical Data Science. 2022;5:293–320. doi: 10.1146/annurev-biodatasci-111721-074830

6. Ruan Y, Lin Y-F, Feng Y-CA, Chen C-Y, Lam M, Guo Z, Ahn YM, Akiyama K, Arai M, Baek JH, et al. Improving polygenic prediction in ancestrally diverse populations. Nature Genetics. 2022;54:573–580. doi: 10.1038/s41588-022-01054-7

7. Norland K, Schaid DJ, Kullo IJ. A linear weighted combination of polygenic scores for a broad range of traits improves prediction of coronary heart disease. European Journal of Human Genetics. 2023. doi: 10.1038/s41431-023-01463-0

8. Bick AG, Metcalf GA, Mayo KR, Lichtenstein L, Rura S, Carroll RJ, Musick A, Linder JE, Jordan IK, Nagar SD, et al. Genomic data in the All of Us Research Program. Nature. 2024. doi: 10.1038/s41586-023-06957-x

9. Mayo KR, Basford MA, Carroll RJ, Dillon M, Fullen H, Leung J, Master H, Rura S, Sulieman L, Kennedy N, et al. The *All of Us* Data and Research Center: Creating a Secure, Scalable, and Sustainable Ecosystem for Biomedical Research. Annual Review of Biomedical Data Science. 2023;6:443–464. doi: 10.1146/annurev-biodatasci-122120-104825

10. Martin AR, Kanai M, Kamatani Y, Okada Y, Neale BM, Daly MJ. Clinical use of current polygenic risk scores may exacerbate health disparities. Nature Genetics. 2019;51:584–591. doi: 10.1038/s41588-019-0379-x

11. Newton KM, Peissig PL, Kho AN, Bielinski SJ, Berg RL, Choudhary V, Basford M, Chute CG, Kullo IJ, Li R, et al. Validation of electronic medical record-based phenotyping algorithms: results and lessons learned from the eMERGE network. Journal of the American Medical Informatics Association. 2013;20:e147-e154. doi: 10.1136/amiajnl-2012-000896

12. Dikilitas O, Schaid DJ, Kosel ML, Carroll RJ, Chute CG, Denny JC, Fedotov A, Feng Q, Hakonarson H, Jarvik GP, et al. Predictive Utility of Polygenic Risk Scores for Coronary Heart Disease in Three Major Racial and Ethnic Groups. The American Journal of Human Genetics. 2020;106:707–716. doi: 10.1016/j.ajhg.2020.04.002

13. Patel AP, Wang M, Ruan Y, Koyama S, Clarke SL, Yang X, Tcheandjieu C, Agrawal S, Fahed AC, Ellinor PT, et al. A multi-ancestry polygenic risk score improves risk prediction for coronary artery disease. Nature Medicine. 2023;29:1793–1803. doi: 10.1038/s41591-023-02429-x

14. Smith JL, Tcheandjieu C, Dikilitas O, Iyer K, Miyazawa K, Hilliard A, Lynch J, Rotter JI, Chen Y-DI, Sheu WH-H, et al. Multi-Ancestry Polygenic Risk Score for Coronary Heart Disease Based on an Ancestrally Diverse Genome-Wide Association Study and Population-Specific Optimization. Circulation: Genomic and Precision Medicine. 2024;0:e004272. doi: doi:10.1161/CIRCGEN.123.004272

15. Lennon NJ, Kottyan LC, Kachulis C, Abul-Husn NS, Arias J, Belbin G, Below JE, Berndt SI, Chung WK, Cimino JJ, et al. Selection, optimization and validation of ten chronic disease polygenic risk scores for clinical implementation in diverse US populations. Nature Medicine. 2024;30:480–487. doi: 10.1038/s41591-024-02796-z

16. Privé F, Aschard H, Ziyatdinov A, Blum MGB. Efficient analysis of large-scale genome-wide data with two R packages: bigstatsr and bigsnpr. Bioinformatics. 2018;34:2781–2787. doi: 10.1093/bioinformatics/bty185

17. Chang CC, Chow CC, Tellier LC, Vattikuti S, Purcell SM, Lee JJ. Second-generation PLINK: rising to the challenge of larger and richer datasets. GigaScience. 2015;4:7. doi: 10.1186/s13742-015-0047-8

18. Wand H, Lambert SA, Tamburro C, Iacocca MA, O’Sullivan JW, Sillari C, Kullo IJ, Rowley R, Dron JS, Brockman D, et al. Improving reporting standards for polygenic scores in risk prediction studies. Nature. 2021;591:211–219. doi: 10.1038/s41586-021-03243-6

19. Tay JK, Narasimhan B, Hastie T. Elastic Net Regularization Paths for All Generalized Linear Models. Journal of Statistical Software. 2023;106. doi: 10.18637/jss.v106.i01

20. Goff DC, Lloyd-Jones DM, Bennett G, Coady S, D’Agostino RB, Gibbons R, Greenland P, Lackland DT, Levy D, O’Donnell CJ, et al. 2013 ACC/AHA Guideline on the Assessment of Cardiovascular Risk. Circulation. 2014;129:S49-S73. doi: 10.1161/01.cir.0000437741.48606.98

21. Privé F, Aschard H, Carmi S, Folkersen L, Hoggart C, O’Reilly PF, Vilhjálmsson BJ. Portability of 245 polygenic scores when derived from the UK Biobank and applied to 9 ancestry groups from the same cohort. The American Journal of Human Genetics. 2022;109:12–23. doi: 10.1016/j.ajhg.2021.11.008

22. Wang Y, Namba S, Lopera E, Kerminen S, Tsuo K, Lall K, Kanai M, Zhou W, Wu K-HH, Fave M-J, et al. Global Biobank analyses provide lessons for developing polygenic risk scores across diverse cohorts. Cell Genomics. 2023;3. doi: 10.1016/j.xgen.2022.100241

23. Roychowdhury T, Klarin D, Levin MG, Spin JM, Rhee YH, Deng A, Headley CA, Tsao NL, Gellatly C, Zuber V, et al. Genome-wide association meta-analysis identifies risk loci for abdominal aortic aneurysm and highlights PCSK9 as a therapeutic target. Nature Genetics. 2023. doi: 10.1038/s41588-023-01510-y

24. Malik R, Chauhan G, Traylor M, Sargurupremraj M, Okada Y, Mishra A, Rutten-Jacobs L, Giese A-K, Van Der Laan SW, Gretarsdottir S, et al. Multiancestry genome-wide association study of 520,000 subjects identifies 32 loci associated with stroke and stroke subtypes. Nature Genetics. 2018;50:524–537. doi: 10.1038/s41588-018-0058-3

25. Dichgans M, Pulit SL, Rosand J. Stroke genetics: discovery, biology, and clinical applications. The Lancet Neurology. 2019;18:587–599. doi: 10.1016/s1474-4422(19)30043-2

26. Vilhjálmsson BJ, Yang J, Finucane HK, Gusev A, Lindström S, Ripke S, Genovese G, Loh P-R, Bhatia G, Do R, et al. Modeling Linkage Disequilibrium Increases Accuracy of Polygenic Risk Scores. The American Journal of Human Genetics. 2015;97:576–592. doi: 10.1016/j.ajhg.2015.09.001

27. Mak TSH, Porsch RM, Choi SW, Zhou X, Sham PC. Polygenic scores via penalized regression on summary statistics. Genetic Epidemiology. 2017;41:469–480. doi: 10.1002/gepi.22050

28. Inouye M, Abraham G, Nelson CP, Wood AM, Sweeting MJ, Dudbridge F, Lai FY, Kaptoge S, Brozynska M, Wang T, et al. Genomic Risk Prediction of Coronary Artery Disease in 480,000 Adults: Implications for Primary Prevention. J American Coll Cardiol. 2018;72:1883–1893.

29. Abraham G, Malik R, Yonova-Doing E, Salim A, Wang T, Danesh J, Butterworth AS, Howson JMM, Inouye M, Dichgans M. Genomic risk score offers predictive performance comparable to clinical risk factors for ischaemic stroke. Nature Communications. 2019;10. doi: 10.1038/s41467-019-13848-1

30. Ge T, Chen C-Y, Ni Y, Feng Y-CA, Smoller JW. Polygenic prediction via Bayesian regression and continuous shrinkage priors. Nature Communications. 2019;10. doi: 10.1038/s41467-019-09718-5

31. Privé F, Arbel J, Vilhjálmsson BJ. LDpred2: better, faster, stronger. Bioinformatics. 2021;36:5424–5431. doi: 10.1093/bioinformatics/btaa1029

32. Cavazos TB, Witte JS. Inclusion of variants discovered from diverse populations improves polygenic risk score transferability. HGG Adv. 2021;2:100017. doi: 10.1016/j.xhgg.2020.100017

33. Tsao CW, Aday AW, Almarzooq ZI, Anderson CAM, Arora P, Avery CL, Baker-Smith CM, Beaton AZ, Boehme AK, Buxton AE, et al. Heart Disease and Stroke Statistics—2023 Update: A Report From the American Heart Association. Circulation. 2023;147:e93-e621. doi: 10.1161/cir.0000000000001123

34. Albiñana C, Zhu Z, Schork AJ, Ingason A, Aschard H, Brikell I, Bulik CM, Petersen LV, Agerbo E, Grove J, et al. Multi-PGS enhances polygenic prediction by combining 937 polygenic scores. Nature Communications. 2023;14. doi: 10.1038/s41467-023-40330-w

35. Truong B, Hull LE, Ruan Y, Huang QQ, Hornsby W, Martin H, Van Heel DA, Wang Y, Martin AR, Lee SH, et al. Integrative polygenic risk score improves the prediction accuracy of complex traits and diseases. Cell Genomics. 2024;4:100523. doi: 10.1016/j.xgen.2024.100523

36. Turley P, Walters RK, Maghzian O, Okbay A, Lee JJ, Fontana MA, Nguyen-Viet TA, Wedow R, Zacher M, Furlotte NA, et al. Multi-trait analysis of genome-wide association summary statistics using MTAG. Nature Genetics. 2018;50:229–237. doi: 10.1038/s41588-017-0009-4

37. Bhattacharjee S, Rajaraman P, Kevin, William, Beatrice, Hartge P, Yeager M, Charles, Stephen, Chatterjee N. A Subset-Based Approach Improves Power and Interpretation for the Combined Analysis of Genetic Association Studies of Heterogeneous Traits. The American Journal of Human Genetics. 2012;90:821–835. doi: 10.1016/j.ajhg.2012.03.015

38. Wang F, Ghanzouri I, Leeper NJ, Tsao PS, Gyang Ross E. Development of a polygenic risk score to improve detection of peripheral artery disease. Vascular Medicine. 2022;27:219–227. doi: 10.1177/1358863X211067564

39. Neumann JT, Riaz M, Bakshi A, Polekhina G, Thao LTP, Nelson MR, Woods RL, Abraham G, Inouye M, Reid CM, et al. Predictive Performance of a Polygenic Risk Score for Incident Ischemic Stroke in a Healthy Older Population. Stroke. 2021;52:2882–2891. doi: 10.1161/strokeaha.120.033670

40. Van Zuydam NR, Stiby A, Abdalla M, Austin E, Dahlström EH, McLachlan S, Vlachopoulou E, Ahlqvist E, Di Liao C, Sandholm N, et al. Genome-Wide Association Study of Peripheral Artery Disease. Circulation: Genomic and Precision Medicine. 2021;14. doi: 10.1161/circgen.119.002862

41. Consortium TCADCDG. A genome-wide association study in Europeans and South Asians identifies five new loci for coronary artery disease. Nature Genetics. 2011;43:339–344. doi: 10.1038/ng.782

42. Tcheandjieu C, Zhu X, Hilliard AT, Clarke SL, Napolioni V, Ma S, Lee KM, Fang H, Chen F, Lu Y, et al. Large-scale genome-wide association study of coronary artery disease in genetically diverse populations. Nature Medicine. 2022;28:1679–1692. doi: 10.1038/s41591-022-01891-3

43. Kullo IJ. Promoting equity in polygenic risk assessment through global collaboration. Nature Genetics. 2024. doi: 10.1038/s41588-024-01843-2

44. Norland K, Schaid DJ, Naderian M, Na J, Kullo IJ. Joint Association of Polygenic Risk and Social Determinants of Health with Coronary Heart Disease in the United States. In: >Cold Spring Harbor Laboratory; 2024.

45. Kullo IJ, Jouni H, Olson JE, Montori VM, Bailey KR. Design of a randomized controlled trial of disclosing genomic risk of coronary heart disease: the Myocardial Infarction Genes (MI-GENES) study. BMC Medical Genomics. 2015;8:51. doi: 10.1186/s12920-015-0122-0

46. Lewis ACF, Chisholm RL, Connolly JJ, Esplin ED, Glessner J, Gordon A, Green RC, Hakonarson H, Harr M, Holm IA, et al. Managing differential performance of polygenic risk scores across groups: Real-world experience of the eMERGE Network. American Journal of Human Genetics. 2024;111:999–1005. doi: 10.1016/j.ajhg.2024.04.005

47. Alexander DH, Lange K. Enhancements to the ADMIXTURE algorithm for individual ancestry estimation. BMC Bioinformatics. 2011;12:246. doi: 10.1186/1471-2105-12-246

48. Chiu AM, Molloy EK, Tan Z, Talwalkar A, Sankararaman S. Inferring population structure in biobank-scale genomic data. The American Journal of Human Genetics. 2022;109:727–737. doi: 10.1016/j.ajhg.2022.02.015

49. Aragam KG, Jiang T, Goel A, Kanoni S, Wolford BN, Atri DS, Weeks EM, Wang M, Hindy G, Zhou W, et al. Discovery and systematic characterization of risk variants and genes for coronary artery disease in over a million participants. Nature Genetics. 2022;54:1803–1815. doi: 10.1038/s41588-022-01233-6

50. Norland K, Schaid DJ, Kullo IJ. A linear weighted combination of polygenic scores for a broad range of traits improves prediction of coronary heart disease. European Journal of Human Genetics. 2023. doi: 10.1038/s41431-023-01463-0

51. Shim I, Kuwahara H, Chen N, Hashem MO, Alabdi L, Abouelhoda M, Won H-H, Natarajan P, Ellinor PT, Khera AV, et al. Clinical utility of polygenic scores for cardiometabolic disease in Arabs. Nature Communications. 2023;14. doi: 10.1038/s41467-023-41985-1

52. Breeyear JH, Shuey MM, Edwards TL, Hellwege JN. Blood Pressure Polygenic Scores Are Associated With Apparent Treatment-Resistant Hypertension. Circulation: Genomic and Precision Medicine. 2022;15. doi: 10.1161/circgen.121.003554

53. Graham SE, Clarke SL, Wu K-HH, Kanoni S, Zajac GJM, Ramdas S, Surakka I, Ntalla I, Vedantam S, Winkler TW, et al. The power of genetic diversity in genome-wide association studies of lipids. Nature. 2021;600:675–679. doi: 10.1038/s41586-021-04064-3

54. Khera AV, Chaffin M, Wade KH, Zahid S, Brancale J, Xia R, Distefano M, Senol-Cosar O, Haas ME, Bick A, et al. Polygenic Prediction of Weight and Obesity Trajectories from Birth to Adulthood. Cell. 2019;177:587–596.e589. doi: 10.1016/j.cell.2019.03.028

