## Supplemental File 1 for "Comparative Performance of Polygenic Risk Scores for Atherosclerotic Cardiovascular Disease Subtypes in the *All of Us* Research Program"

#### Table of Contents


1. Supplemental Figures S1-S11
2. Supplemental Tables S1-S2


#### Supplemental Figures

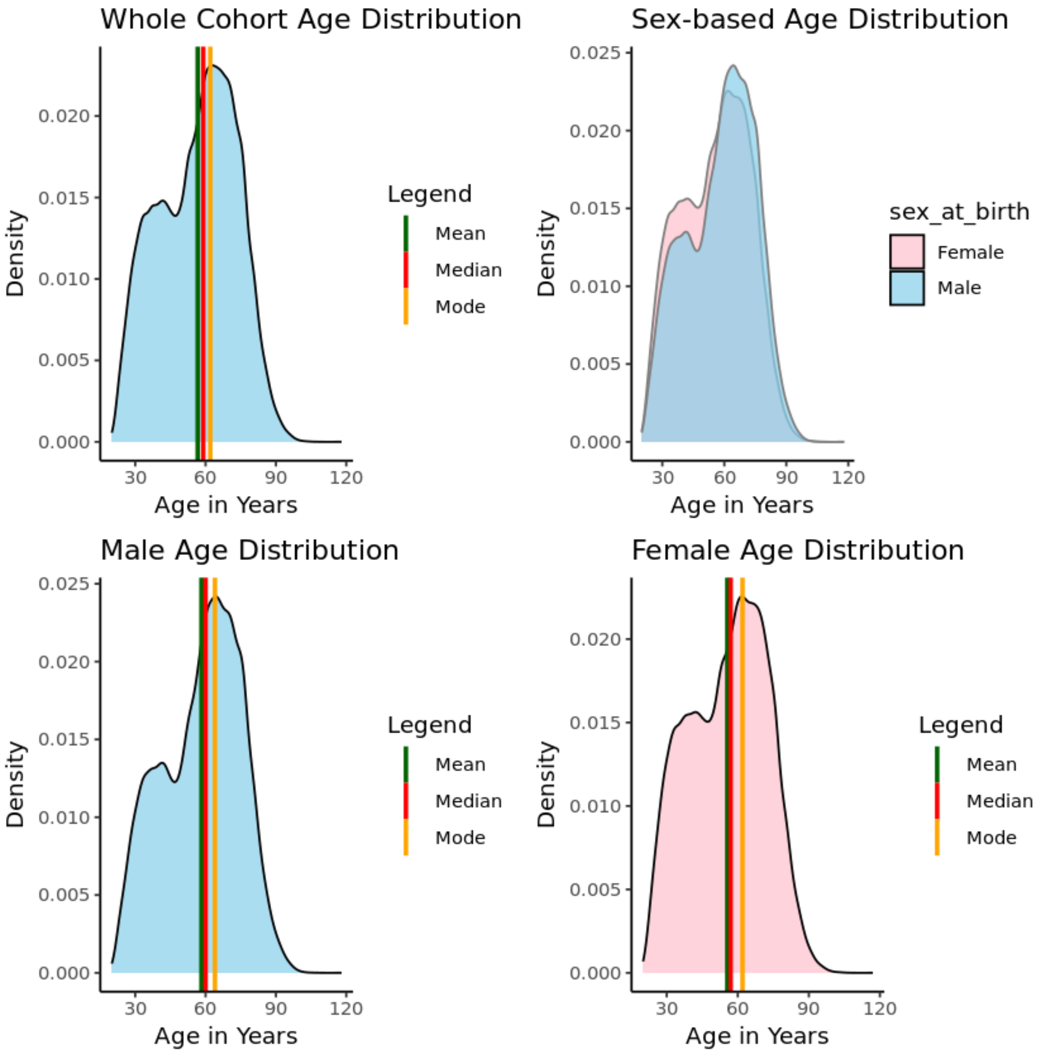


*Figure S1. Age distribution a.) with mean, median, and mode across the whole ASCVD validation cohort, b.) comparing sex at birth designations overlayed, and for c.) male and d.) female individuals in the cohort.*

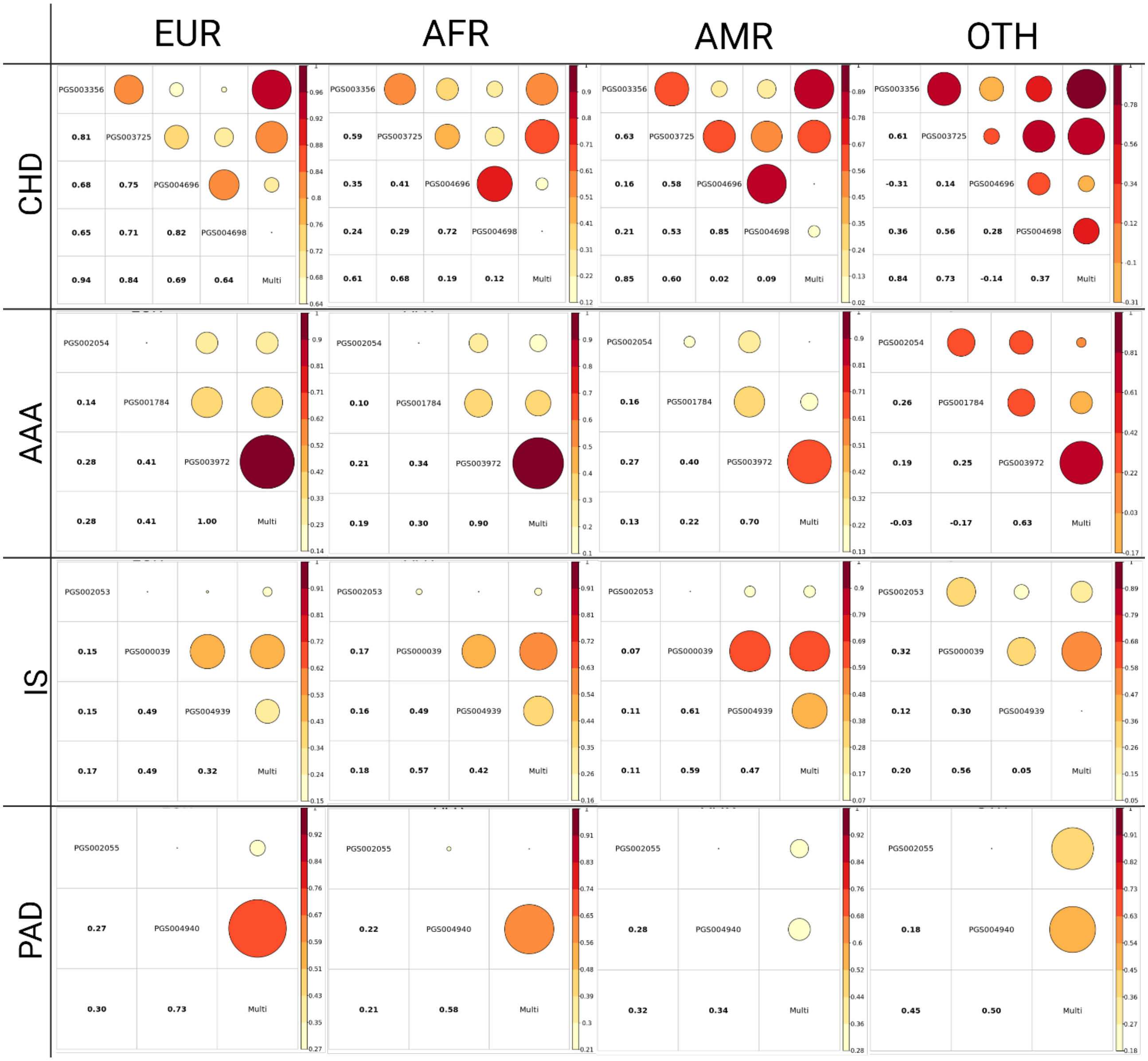


*Figure S2. Correlations of PRS within each phenotype A) CHD, B) IS, C) AAA, and D) PAD for European (EUR), African (AFR), Admixed American (AMR), and Other (OTH) genetic ancestry groups.*

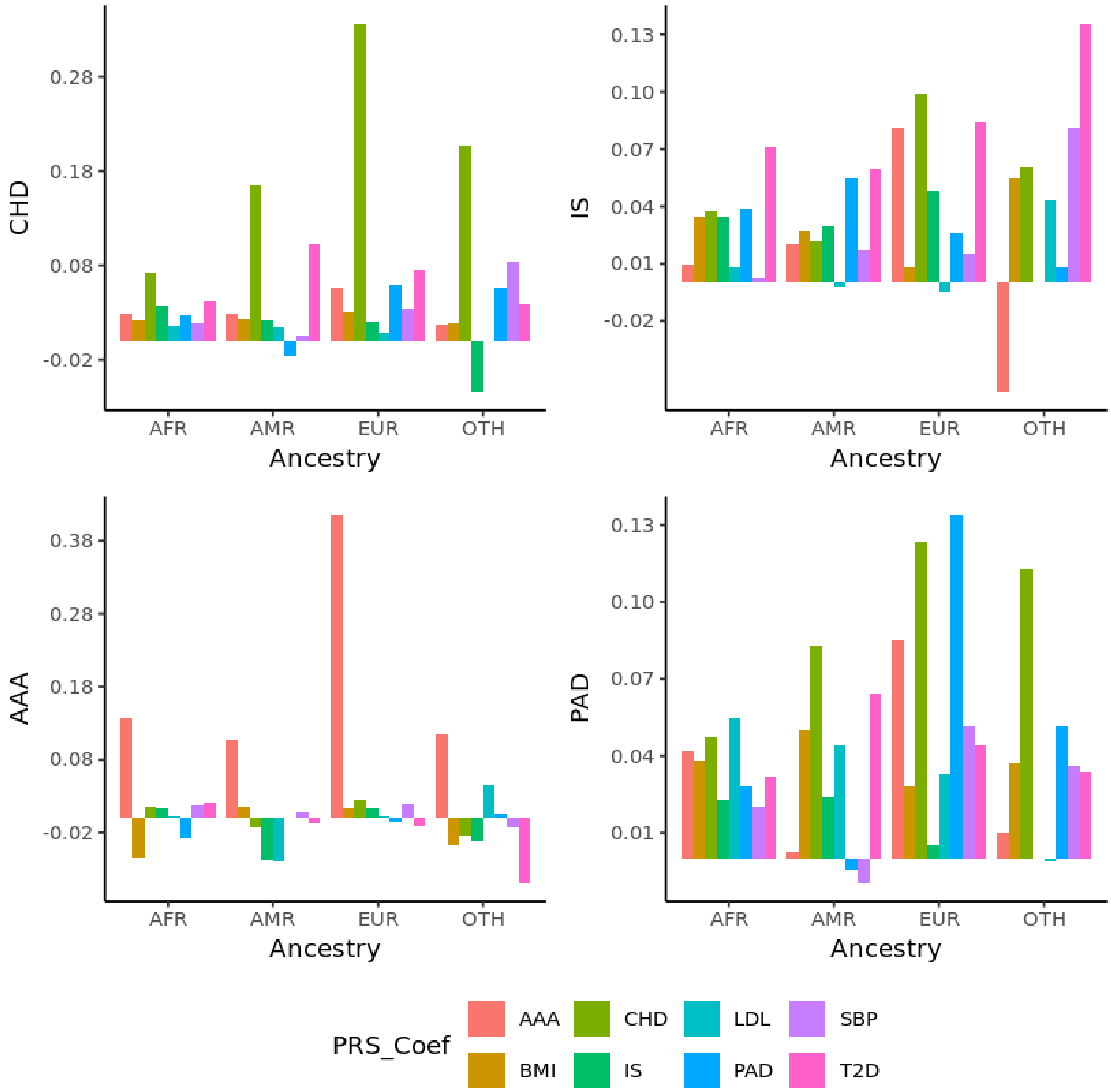


*Figure S3. Coefficients for adjustment of multi-trait PRS for all subtypes with values for all variables used in the PRS development across European (EUR), African (AFR), Admixed American (AMR), and Other (OTH) genetic ancestry groups.*

.

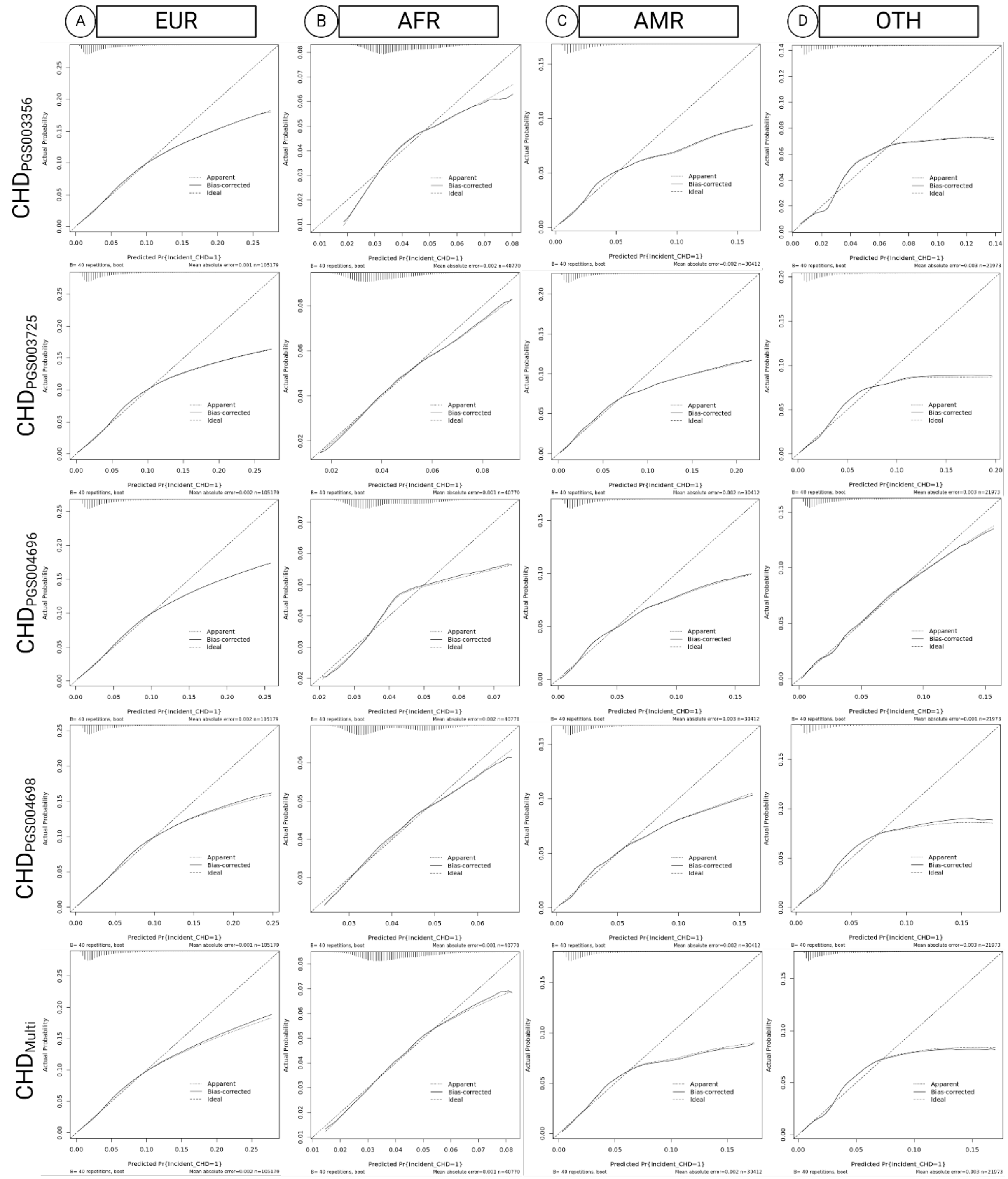


*Figure S4. Calibration of PRS for CHD adjusted for age and sex with predicted probability on the x-axis and actual probability on the y-axis for European (EUR), African (AFR), Admixed American (AMR), and Other (OTH) genetic ancestry groups. Distribution of the cases is shown in the top left corner of each plot.*

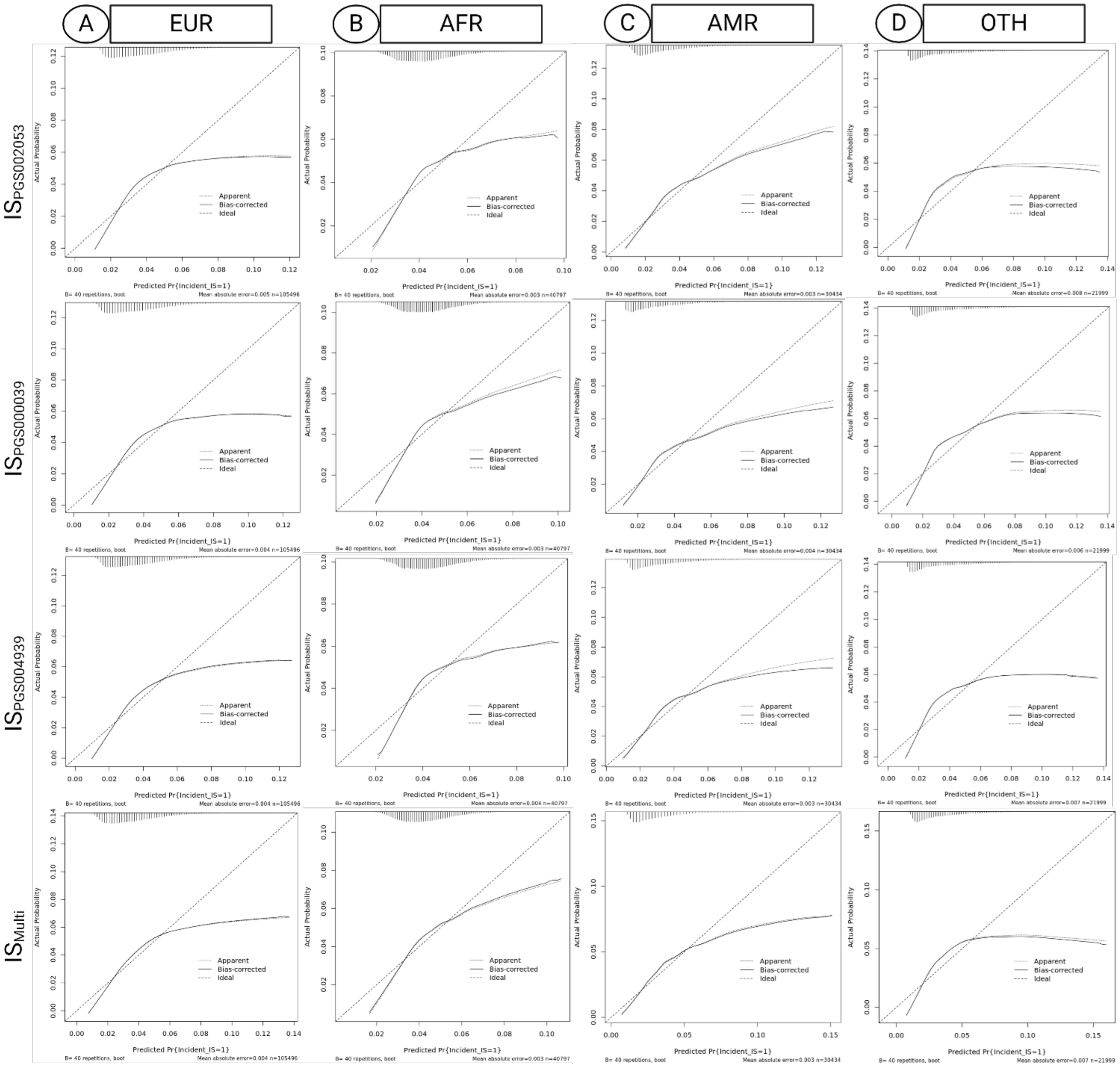


*Figure S5. Calibration of PRS for IS adjusted for age and sex with predicted probability on the x-axis and actual probability on the y-axis for European (EUR), African (AFR), Admixed American (AMR), and Other (OTH) genetic ancestry groups. Distribution of the cases is shown in the top left corner of each plot.*

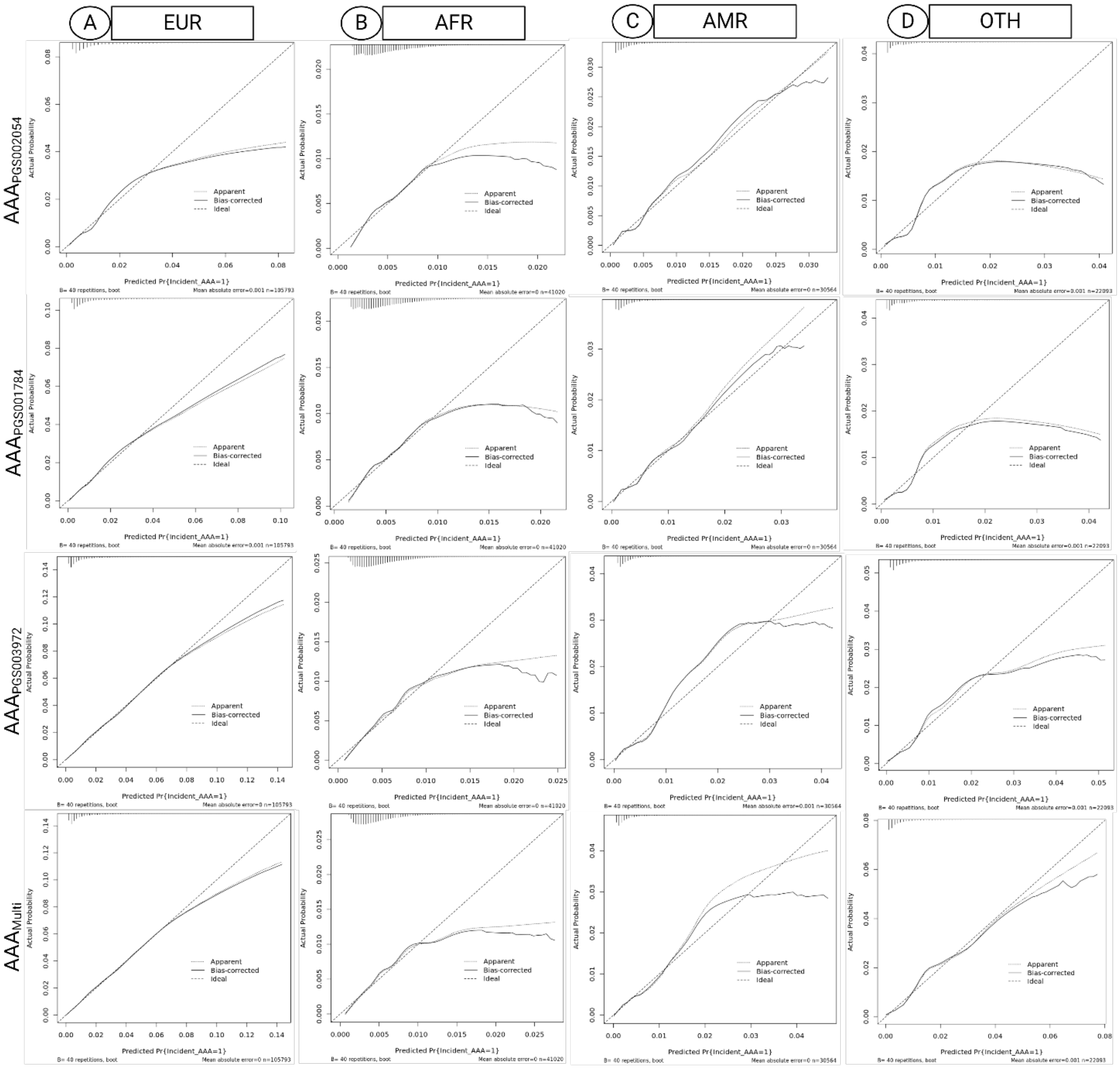


*Figure S6. Calibration of PRS for AAA adjusted for age and sex with predicted probability on the x-axis and actual probability on the y-axis for European (EUR), African (AFR), Admixed American (AMR), and Other (OTH) genetic ancestry groups. Distribution of the cases is shown in the top left corner of each plot.*

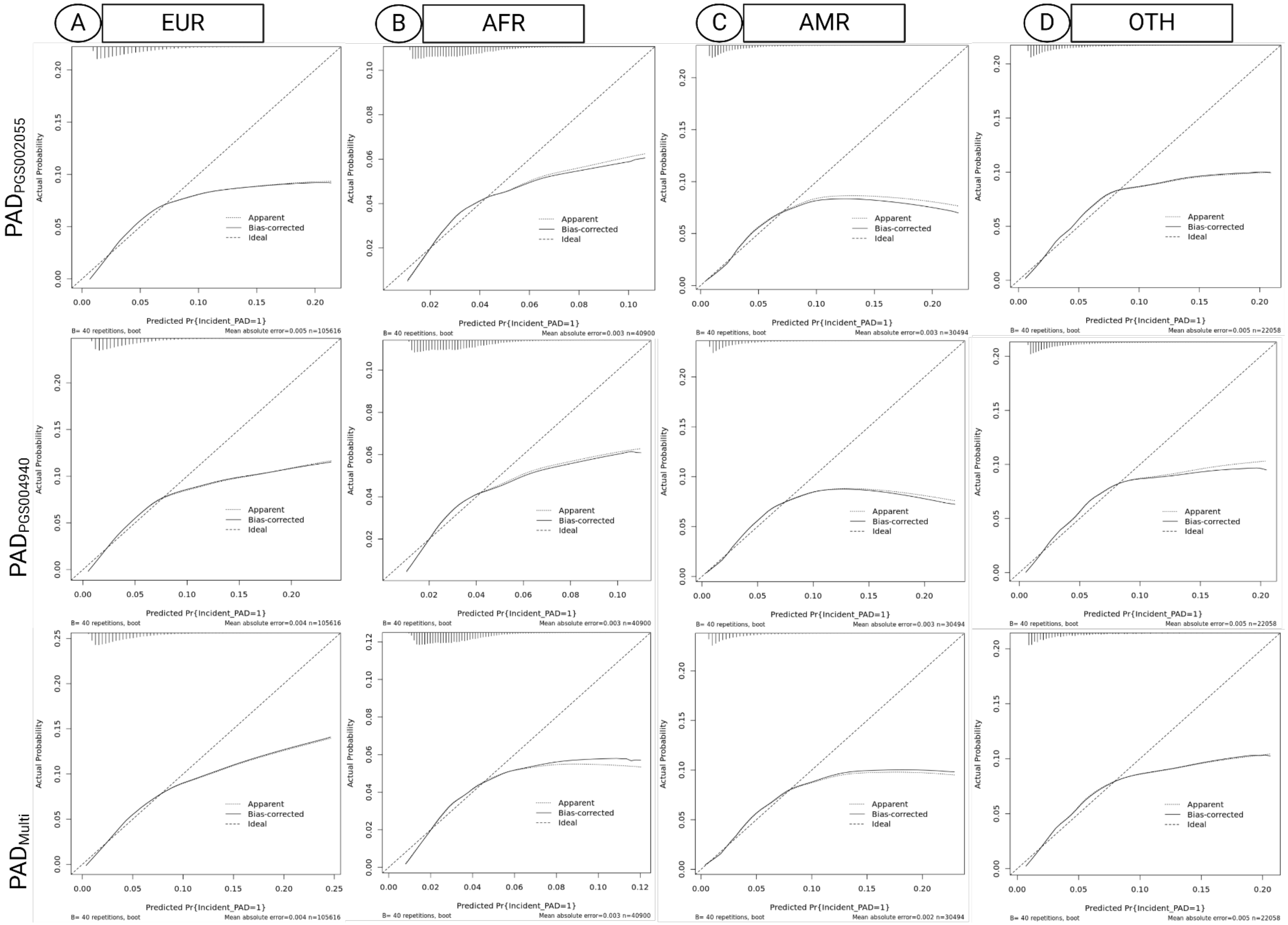


*Figure S7. Calibration of PRS for PAD adjusted for age and sex with predicted probability on the x-axis and actual probability on the y-axis for European (EUR), African (AFR), Admixed American (AMR), and Other (OTH) genetic ancestry groups. Distribution of the cases is shown in the top left corner of each plot.*

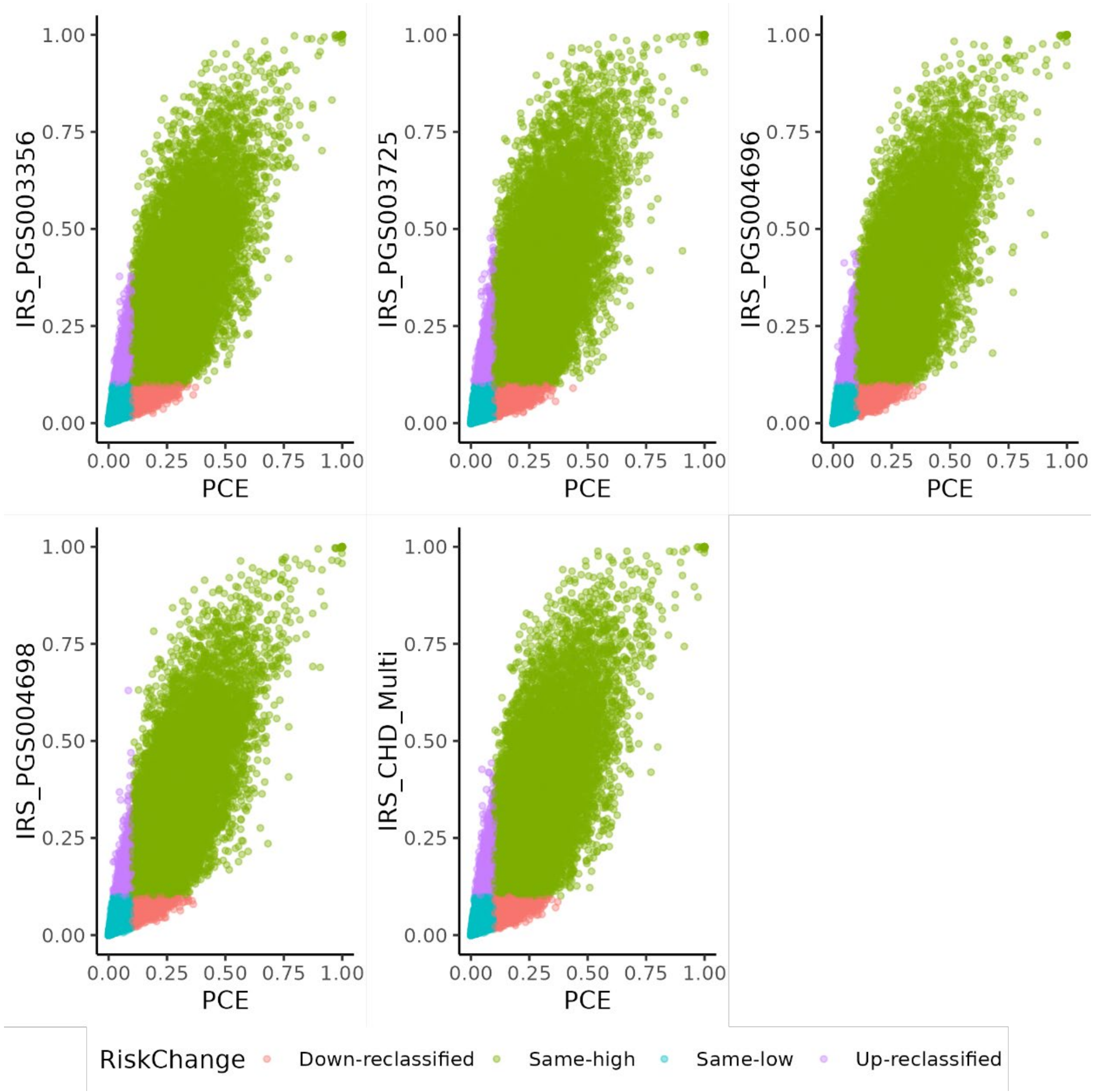


*Figure S8. Net reclassification plots for each IRS based on each PRS for CHD and integration with the clinical PCE for 10-year clinical risk estimates with a threshold of > 10% representing high risk. Blue represents same category low risk, purple represents up-reclassified risk, pink represents down-reclassified risk, and green represents same category high risk.*

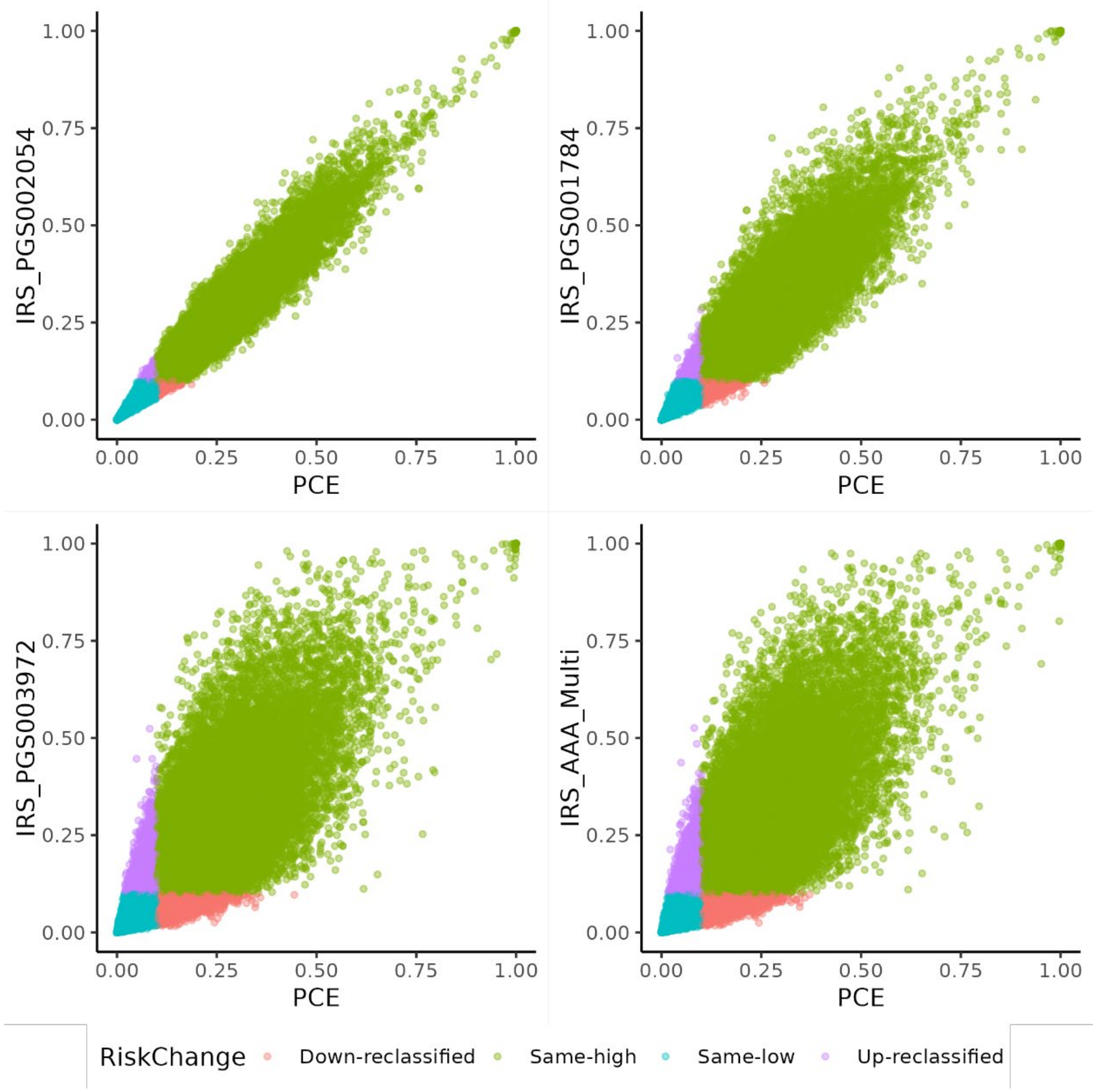


*Figure S9. Net reclassification plots for each IRS based on each PRS for AAA and integration with the clinical PCE for 10-year clinical risk estimates with a threshold of > 10% representing high risk. Blue represents same category low risk, purple represents up-reclassified risk, pink represents down-reclassified risk, and green represents same category high risk.*

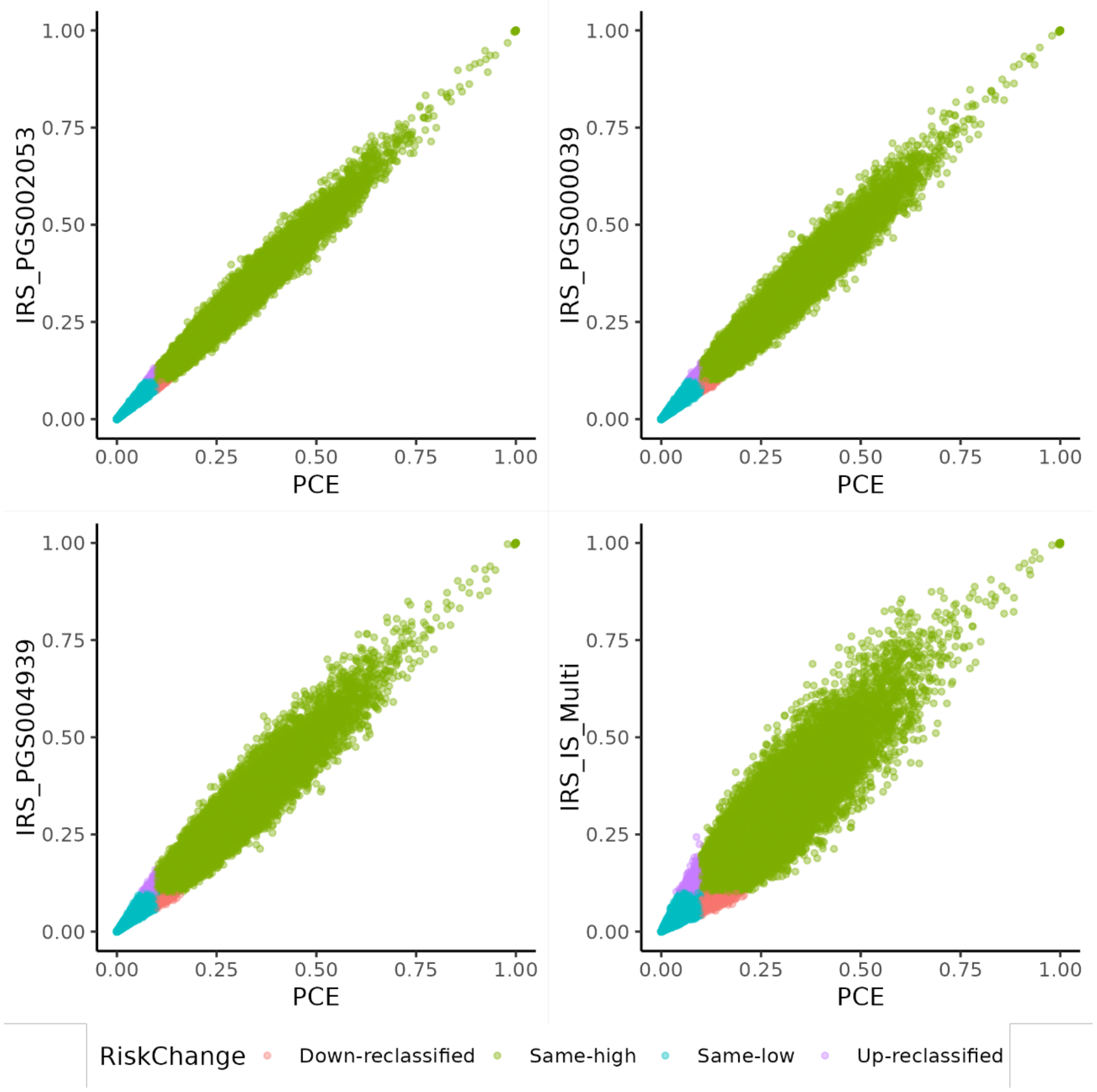


*Figure S10. Net reclassification plots for each IRS based on each PRS for IS and integration with the clinical PCE for 10-year clinical risk estimates with a threshold of > 10% representing high risk. Blue represents same category low risk, purple represents up-reclassified risk, pink represents down-reclassified risk, and green represents same category high risk.*

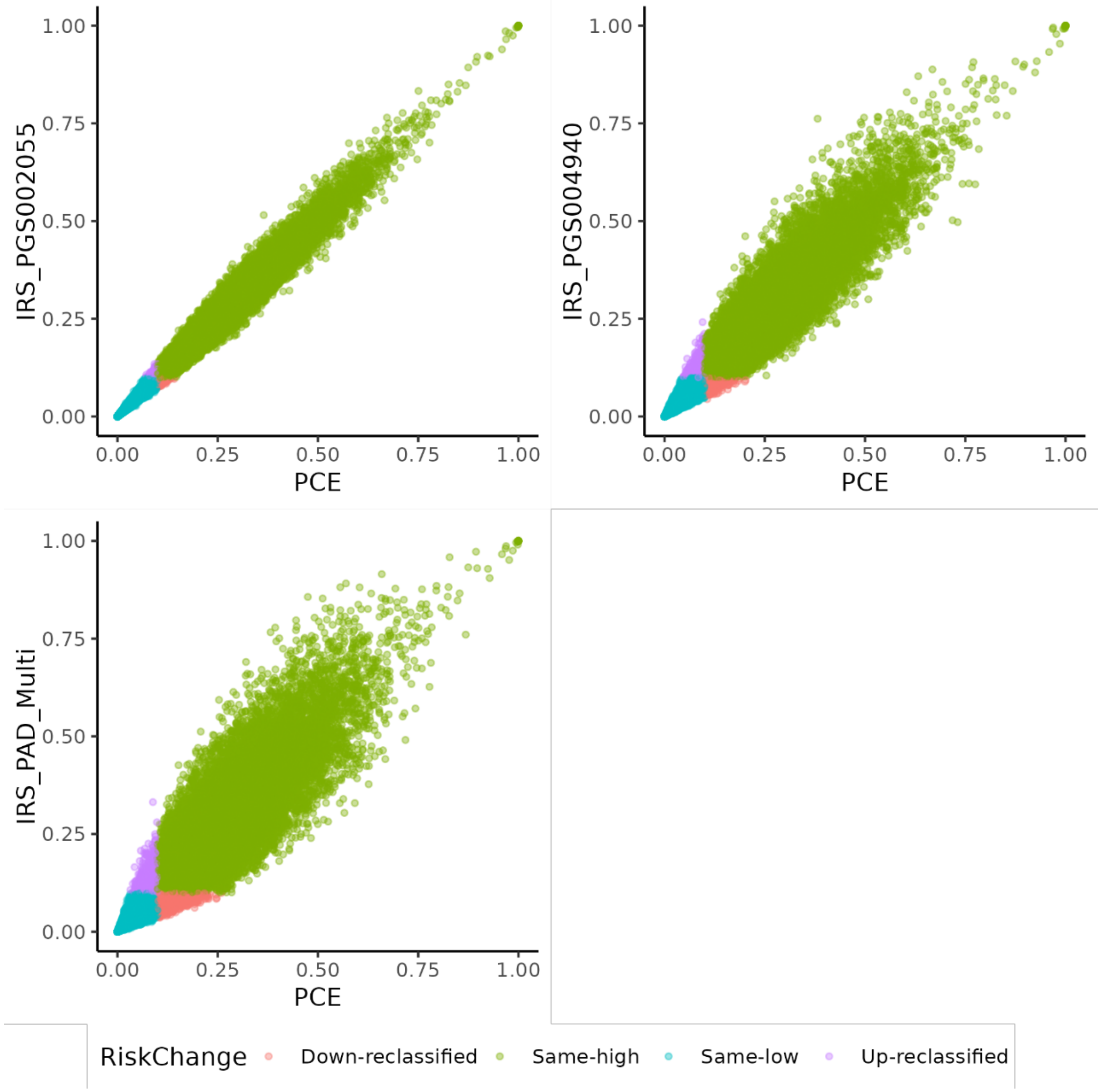


*Figure S11. Net reclassification plots for each IRS based on each PRS for PAD and integration with the clinical PCE for 10-year clinical risk estimates with a threshold of > 10% representing high risk. Blue represents same category low risk, purple represents up-reclassified risk, pink represents down-reclassified risk, and green represents same category high risk.*

### Supplemental Tables

*Table S1. Average years to event for variable ascertainment for ancestry-specific populations (EUR, AFR, AMR) in each phenotype of interest (CHD, IS, AAA, PAD).*

| | CHD[*] | IS[†] | AAA[‡] | PAD[§] |
|---|---|---|---|---|
| EUR[||] | 8 | 8 | 9 | 8 |
| AFR[#] | 8 | 8 | 8 | 8 |
| AMR[**] | 6 | 6 | 7 | 6 |
| OTH[††] | 7 | 7 | 8 | 7 |

[*]CHD- coronary heart disease
[†]IS- ischemic stroke
[‡]AAA- abdominal aortic aneurysm
[§]PAD- peripheral artery disease
[||]EUR- European
[#]AFR- African
[**]AMR- Admixed American
[††]OTH- Other

*Table S2. Performance of PRS for ASCVD phenotypes (CHD, IS, AAA, PAD) adjusted for age and sex, for European (EUR), African (AFR), Admixed American (AMR), and Other (OTH) genetic ancestry groups. Odds ratio (OR) per SD (95% CI), AUC of the model, AUC of the base model, P-values, Brier Scores, and calibration $R^2$ values are provided.*

| **Trait** | **PGSCatalog ID** | **Ancestry** | **Case/Control** | **OR per SD (95% CI)** [*] | **C-Statistic$_{base}$**[†] | **C-Statistic$_{base+PRS}$**[‡] | **P-value** | **Brier Score** | **$R^2$**[§] | **Top 5% vs Rest OR (95% CI)** [||] | **C-Statistic$_{Top5\%}$** [#] | **P-Value** |
|---|---|---|---|---|---|---|---|---|---|---|---|---|
| **CHD** | PGS003725 | EUR | 3,744/101,435 | 1.668 (1.613-1.725) | 0.649 | 0.712 (0.704-0.720) | 2.59E-194 | 0.038 | 0.073 | 2.734 (2.464-3.034) | 0.667 | 6.99E-80 |
| | | AFR | 1,604/39,116 | 1.215 (1.155-1.278) | 0.553 | 0.582 (0.568-0.596) | 3.71E-14 | 0.034 | 0.010 | 1.512 (1.246-1.834) | 0.559 | 2.78E-05 |
| | | AMR | 694/29,718 | 1.539 (1.427-1.661) | 0.691 | 0.691 (0.702-0.738) | 1.01E-28 | 0.022 | 0.066 | 2.225 (1.730-2.862) | 0.697 | 4.80E-10 |
| | | OTH | 573/21,400 | 1.528 (1.402-1.666) | 0.721 | 0.750 (0.732-0.768) | 1.93E-262 | 0.025 | 0.079 | 2.817 (2.112-3.756) | 0.733 | 1.74E-12 |

| | | | | | | | | | | | | |
|---|---|---|---|---|---|---|---|---|---|---|---|---|
| | PGS004696 | EUR | | 1.616 (1.563-1.671) | | 0.706 (0.698-0.714) | 9.35E-177 | 0.038 | 0.069 | 2.495 (2.242-2.777) | 0.664 | 3.85E-63 |
| | | AFR | | 1.125 (1.071-1.183) | | 0.570 (0.556-0.584) | 3.34E-06 | 0.034 | 0.007 | 1.453 (1.195-1.768) | 0.558 | 1.85E-04 |
| | | AMR | | 1.431 (1.322-1.550) | | 0.710 (0.692-0.728) | 1.12E-18 | 0.022 | 0.059 | 1.976 (1.534-2.545) | 0.698 | 1.35E-07 |
| | | OTH | | 1.609 (1.441-1.797) | | 0.739 (0.719-0.759) | 2.99E-17 | 0.025 | 0.084 | 2.403 (1.860-3.105) | 0.731 | 1.98E-11 |
| | PGS004698 | EUR | | 1.558 (1.507-1.610) | | 0.700 (0.692-0.708) | 5.03E-150 | 0.038 | 0.066 | 2.161 (1.931-2.419) | 0.660 | 6.06E-41 |
| | | AFR | | 1.090 (1.037-1.145) | | 0.564 (0.550-0.578) | 6.77E-04 | 0.034 | 0.006 | 1.281 (1.043-1.574) | 0.555 | 1.83E-02 |
| | | AMR | | 1.448 (1.338-1.567) | | 0.712 (0.694-0.730 ) | 3.46E-20 | 0.022 | 0.059 | 1.838 (1.416-2.387) | 0.697 | 4.90E-06 |
| | | OTH | | 1.385 (1.271-1.510) | | 0.737 (0.717-0.757) | 1.05E-13 | 0.025 | 0.073 | 1.816 (1.302-2.532) | 0.724 | 4.43E-04 |
| | PGS003356 | EUR | | 1.612 (1.559-1.667) | | 0.705 (0.697-0.713) | 1.19E-172 | 0.038 | 0.070 | 2.666 (2.401-2.960) | 0.666 | 1.70E-75 |
| | | AFR | | 1.159 (1.102-1.218) | | 0.574 (0.560-0.588) | 8.01E-09 | 0.034 | 0.008 | 1.313 (1.070-1.612) | 0.556 | 9.16E-03 |
| | | AMR | | 1.321 (1.225-1.426) | | 0.704 (0.686-0.722) | 6.26E-13 | 0.022 | 0.053 | 1.564 (1.161-2.105) | 0.693 | 3.23E-03 |
| | | OTH | | 1.196 (1.098-1.302) | | 0.726 (0.706-0.746) | 3.76E-05 | 0.025 | 0.063 | 1.148 (0.767-1.717) | 0.721 | 5.03E-01 |

| | | | | | | | | | | | | |
|---|---|---|---|---|---|---|---|---|---|---|---|---|
| | Multi | EUR | | 1.630 (1.576-1.686) | | 0.708 (0.700-0.716) | 4.80E-180 | 0.034 | 0.071 | 2.720 (2.450-3.020) | 0.666 | 2.42E-78 |
| | | AFR | | 1.203 (1.143-1.266) | | 0.580 (0.566-0.594) | 1.49E-12 | 0.038 | 0.009 | 1.473 (1.210-1.792) | 0.558 | 1.10E-04 |
| | | AMR | | 1.409 (1.306-1.521) | | 0.710 (0.692-0.728) | 1.60E-18 | 0.022 | 0.057 | 1.976 (1.534-2.545) | 0.694 | 1.35E-07 |
| | | OTH | | 1.341 (1.233-1.460) | | 0.735 (0.715-0.755) | 9.85E-12 | 0.025 | 0.071 | 2.403 (1.860-3.105) | 0.726 | 1.98E-11 |
| **AAA** | PGS002054 | EUR | 1,161/104,632 | 1.185 (1.118-1.256) | 0.744 | 0.751 (0.739-0.763) | 1.14E-08 | 0.011 | 0.076 | 1.299 (1.022-1.651) | 0.745 | 3.28E-02 |
| | | AFR | 196/40,824 | 0.959 (0.834-1.104) | 0.696 | 0.696 (0.663-0.729) | 5.63E-01 | 0.005 | 0.021 | 0.716 (0.336-1.525) | 0.696 | 3.86E-01 |
| | | AMR | 88/30,476 | 1.076 (0.873-1.326) | 0.773 | 0.772 (0.727-0.817) | 4.94E-01 | 0.003 | 0.051 | 0.973 (0.356-2.663) | 0.773 | 9.58E-01 |
| | | OTH | 123/21,970 | 1.044 (0.873-1.247) | 0.775 | 0.775 (0.740-0.810) | 6.9E-01 | 0.006 | 0.068 | 1.142 (0.500-2.608) | 0.776 | 7.53E-01 |
| | PGS001784 | EUR | | 1.335 (1.260-1.415) | | 0.760 (0.748-0.772) | 1.59E-22 | 0.011 | 0.081 | 2.289 (1.887-2.775) | 0.751 | 4.06E-17 |
| | | AFR | | 1.032 (0.897-1.187) | | 0.696 (0.663-0.729) | 6.62E-01 | 0.005 | 0.021 | 1.135 (0.616-2.090) | 0.696 | 6.85E-01 |
| | | AMR | | 1.178 (0.953-1.455) | | 0.773 (0.728-0.818) | 1.29E-01 | 0.003 | 0.046 | 1.915 (0.880-4.166) | 0.774 | 1.01E-01 |
| | | OTH | | 0.960 (0.797-1.156) | | 0.775 (0.740-0.810) | 6.65E-01 | 0.006 | 0.076 | 1.358 (0.593-3.114) | 0.776 | 4.69E-01 |

| | | | | | | | | | | | | |
|---|---|---|---|---|---|---|---|---|---|---|---|---|
| | PGS003972 | EUR | | 1.631 (1.538-1.730) | | 0.778 (0.766-0.790) | 1.37E-59 | 0.011 | 0.093 | 2.918 (2.436-3.496) | 0.755 | 3.27E-31 |
| | | AFR | | 1.237 (1.073-1.426) | | 0.704 (0.671-0.737) | 3.38E-03 | 0.005 | 0.023 | 1.846 (1.119-3.045) | 0.699 | 1.64E-02 |
| | | AMR | | 1.331 (1.083-1.636) | | 0.776 (0.731-0.821) | 6.64E-03 | 0.003 | 0.060 | 1.520 (0.661-3.496) | 0.772 | 3.25E-01 |
| | | OTH | | 1.426 (1.181-1.723) | | 0.789 (0.754-0.824) | 2.34E-04 | 0.006 | 0.077 | 1.676 (0.812-3.460) | 0.777 | 1.62E-01 |
| | Multi | EUR | | 1.633 (1.540-1.733) | | 0.778 (0.766-0.790) | 7.26E-60 | 0.011 | 0.094 | 2.293 (1.891-2.781) | 0.751 | 3.40E-17 |
| | | AFR | | 1.306 (1.134-1.504) | | 0.710 (0.677-0.743) | 2.14E-04 | 0.005 | 0.026 | 1.094 (0.594-2.015) | 0.696 | 7.73E-01 |
| | | AMR | | 1.522 (1.240-1.869) | | 0.790 (0.749-0.831) | 5.96E-05 | 0.003 | 0.063 | 1.776 (0.771-4.088) | 0.772 | 1.77E-01 |
| | | OTH | | 1.681 (1.370-2.063) | | 0.795 (0.756-0.834) | 6.53E-07 | 0.006 | 0.089 | 1.245 (0.603-2.572) | 0.776 | 5.53E-01 |
| **IS** | PGS002053 | EUR | 3,720/101,776 | 1.085 (1.050-1.121) | 0.621 | 0.624 (0.616-0.632) | 1.01E-06 | 0.034 | 0.025 | 1.103 (0.954-1.276) | 0.621 | 1.85E-01 |
| | | AFR | 1,732/39,065 | 1.052 (1.002-1.103) | 0.593 | 0.595 (0.583-0.607) | 4.12E-02 | 0.041 | 0.009 | 1.211 (0.987-1.486) | 0.594 | 6.71E-02 |
| | | AMR | 904/29,530 | 1.153 (1.079-1.232) | 0.685 | 0.688 (0.672-0.704) | 2.74E-05 | 0.029 | 0.032 | 1.485 (1.145-1.925) | 0.686 | 2.89E-03 |
| | | OTH | 729/21,270 | 1.005 (0.932-1.083) | 0.695 | 0.695 (0.679-0.711) | 9.04E-01 | 0.032 | 0.040 | 0.912 (0.628-1.325) | 0.695 | 6.30E-01 |

| | | | | | | | | | | | | |
|---|---|---|---|---|---|---|---|---|---|---|---|---|
| | PGS000039 | EUR | | 1.116 (1.080-1.153) | | 0.626 (0.618-0.634) | 5.13E-11 | 0.034 | 0.025 | 1.242 (1.081-1.428) | 0.622 | 2.22E-03 |
| | | AFR | | 1.080 (1.029-1.134) | | 0.596 (0.584-0.608) | 1.17E-03 | 0.041 | 0.010 | 1.191 (0.967-1.468) | 0.594 | 9.97E-02 |
| | | AMR | | 1.032 (0.965-1.103) | | 0.685 (0.669-0.701) | 3.65E-01 | 0.029 | 0.030 | 1.036 (0.755-1.422) | 0.685 | 8.24E-01 |
| | | OTH | | 0.980 (0.905-1.061) | | 0.696 (0.680-0.712) | 6.14E-01 | 0.032 | 0.041 | 0.552 (0.334-0.912) | 0.697 | 2.03E-02 |
| | PGS004939 | EUR | | 1.143 (1.106-1.181) | | 0.628 (0.620-0.636) | 1.54E-15 | 0.034 | 0.027 | 1.361 (1.190-1.556) | 0.623 | 6.84E-06 |
| | | AFR | | 1.059 (1.010-1.111) | | 0.596 (0.584-0.608) | 1.95E-02 | 0.041 | 0.009 | 1.043 (0.837-1.300) | 0.593 | 7.06E-01 |
| | | AMR | | 1.103 (1.032-1.179) | | 0.687 (0.671-0.703) | 3.91E-03 | 0.029 | 0.030 | 1.152 (0.856-1.551) | 0.686 | 3.51E-01 |
| | | OTH | | 1.006 (0.932-1.087) | | 0.695 (0.679-0.711) | 8.71E-01 | 0.032 | 0.040 | 1.045 (0.727-1.502) | 0.695 | 8.12E-01 |
| | Multi | EUR | | 1.239 (1.199-1.281) | | 0.638 (0.630-0.646) | 6.17E-37 | 0.003 | 0.029 | 1.633 (1.437-1.855) | 0.626 | 5.00E-14 |
| | | AFR | | 1.144 (1.090-1.020) | | 0.603 (0.591-0.615) | 6.80E-08 | 0.040 | 0.011 | 1.473 (1.216-1.784) | 0.599 | 7.51E-05 |
| | | AMR | | 1.222 (1.142-1.307) | | 0.692 (0.676-0.708) | 5.38E-09 | 0.029 | 0.033 | 1.523 (1.160-1.998) | 0.686 | 2.43E-03 |
| | | OTH | | 1.293 (1.200-1.394) | | 0.708 (0.692-0.724) | 1.88E-11 | 0.032 | 0.045 | 2.149 (1.623-2.846) | 0.701 | 9.09E-08 |

| | | | | | | | | | | | | |
|---|---|---|---|---|---|---|---|---|---|---|---|---|
| **PAD** | PGS002055 | EUR | 3,982/101,634 | 1.100 (1.065-1.136) | 0.671 | 0.673 (0.665-681) | 4.75E-09 | 0.036 | 0.054 | 1.212 (1.057-1.391) | 0.671 | 6.01E-03 |
| | | AFR | 1,205/39,695 | 1.035 (0.977-1.096) | 0.642 | 0.642 (0.628-0.656) | 2.40E-01 | 0.028 | 0.024 | 1.046 (0.806-1.357) | 0.642 | 7.36E-01 |
| | | AMR | 798/29,696 | 1.071 (0.977-1.150) | 0.725 | 0.726 (0.710-0.742) | 5.99E-02 | 0.025 | 0.068 | 0.982 (0.698-1.382) | 0.725 | 9.16E-01 |
| | | OTH | 738/21,320 | 0.961 (0.890-1.037) | 0.745 | 0.745 (0.729-0.761) | 3.02E-01 | 0.032 | 0.075 | 0.747 (0.480-1.164) | 0.745 | 1.97E-01 |
| | PGS004940 | EUR | | 1.227 (1.189-1.266) | | 0.689 (0.681-0.697) | 5.76E-37 | 0.036 | 0.059 | 1.969 (1.752-2.213) | 0.677 | 6.96E-30 |
| | | AFR | | 1.058 (0.998-1.120) | | 0.648 (0.634-0.662) | 5.63E-02 | 0.028 | 0.024 | 1.696 (1.363-2.111) | 0.647 | 2.20E-06 |
| | | AMR | | 1.103 (1.028-1.184) | | 0.730 (0.714-0.746) | 6.23E-03 | 0.025 | 0.068 | 1.615 (1.209-2.159) | 0.727 | 1.19E-03 |
| | | OTH | | 1.104 (1.020-1.195) | | 0.745 (0.729-0.761) | 1.45E-02 | 0.032 | 0.075 | 1.112 (0.759-1.630) | 0.745 | 5.86E-01 |
| | Multi | EUR | | 1.332 (1.290-1.375) | | 0.676 (0.668-0.684) | 9.53E-57 | 0.036 | 0.063 | 1.396 (1.224-1.592) | 0.672 | 6.33E-07 |
| | | AFR | | 1.147 (1.082-1.216) | | 0.643 (0.629-0.657) | 3.77E-06 | 0.028 | 0.026 | 1.091 (0.845-1.408) | 0.642 | 5.08E-01 |
| | | AMR | | 1.213 (1.129-1.303) | | 0.726 (0.71-0.742) | 1.39E-07 | 0.025 | 0.071 | 0.982 (0.698-1.382) | 0.726 | 9.16E-01 |
| | | OTH | | 1.088 (1.008-1.174) | | 0.745 (0.729-0.761) | 3.12E-02 | 0.032 | 0.076 | 0.747 (0.480-1.163) | 0.745 | 1.97E-01 |

*OR per SD (95% CI)- Odds ratio per standard deviation with 95% confidence interval

[†]C-Statistic$_{base}$ - Concordance statistic of the base model consisting of age and sex at birth
[‡]C-Statistic$_{base+PRS}$ - Concordance statistic of the base model and PRS
[§]$R^2$- Calibration $R^2$
[||]Top 5% vs Rest OR (95% CI)- Odds ratio for the top 5% of risk distribution vs the other 95% of the base model and PRS
[#]C-Statistic$_{Top5\%}$- Concordance statistic for the top 5% of risk distribution vs the other 95% of the base model and PRS
