## Supplemental File 2 for "Comparative Performance of Polygenic Risk Scores for Atherosclerotic Cardiovascular Disease Subtypes in the *All of Us* Research Program"

### Table Key: A description of all columns in all tables.

**Table S3. Original PRS Performance**

| | |
|---|---|
| Trait | ASCVD Subtype |
| PRS | Polygenic risk score PGS Catalog ID |
| Ancestry Tested | Target population for PRS weights |
| HR/OR(95%) | Hazard Ratio or Odds Ratio per standard deviation with 95% confidence intervals when available |

**Table S4. Integrated Risk Score Performance**

| | |
|---|---|
| Prediction Model | Integrated risk score model including clinical risk score and PGS used |
| AUC_CI | Area under the curve with 95% confidence interval for each integrated risk score |
| n_total | number of total individuals used for each analysis |
| Sen_10 | sensitivity of the analysis with a 10% risk threshold |
| Spe_10 | Specificity of the analysis with a 10% risk threshold |
| Accur_10 | Accuracy of the analysis with a 10% risk threshold |
| net_benefit_10 | Net benefit of the analysis with a 10% risk threshold |
| NRI_10 | Net reclassification index using categorical analysis of 10% risk threshold |
| lowerci_10cat | Lower confidence interval of the categorical NRI analysis with 10% risk threshold |
| upperci_10cat | Upper confidence interval of the categorical NRI analysis with 10% risk threshold |
| Cont_NRI | Net reclassification index using continuous analysis |
| lowerci_cont | Lower confidence interval of the continuous NRI |
| upperci_cont | Upper confidence interval of the continuous NRI |

**Table S3. Original PRS Performance**

| **Trait** | **PRS** | **Ancestry tested** | **HR/OR(95%)** |
|---|---|---|---|
| **AAA** | PGS001784 | All | in decile comparison |
| | PGS002054 | All | partial correlation as the performance metric |
| | PGS003972 | All | in decile comparison |
| **CHD** | PGS004696 | EUR | 1.65 (1.59-1.71) |
| | | AFR | 1.20 (1.15-1.26) |
| | | HIS | 1.51 (1.35-1.69) |
| | | EAS | 1.59 (1.54-1.64) |
| | | SAS | 2.67 (2.38-3.00) |
| | PGS004698 | EUR | 1.65 (1.59-1.72) |
| | | AFR | 1.16 (1.11-1.21) |
| | | HIS | 1.38 (1.24-1.54) |
| | | EAS | 1.56 (1.50-1.61) |
| | | SAS | 2.75 (2.41-3.14) |
| | PGS003725 | EUR | 2.14 (2.10-2.19) |
| | | AFR | 1.25 (1.21-1.29) |
| | | HIS | 1.61 (1.53-1.7) |
| | | SAS | 1.83 (1.69-1.99) |
| | PGS003356 | All | 1.563 |
| **IS** | PGS004939 | All | 1.21 (1.19-1.22) |
| | PGS000039 | All | 1.26 (1.22-1.31) |
| | PGS002053 | All | partial correlation as the performance metric |
| **PAD** | PGS004940 | All | 1.28 (1.26-1.30) |
| | PGS002055 | All | partial correlation as the performance metric |

Table S4. Integrated Risk Score Performance

| Prediction_model | AUC_CI | n_total | Sen_10 | Spe_10 | Accur_10 | net_benefit_10 | NRI_10 | lowerci_10cat | upperci_10cat | Cont_NRI | lowerci_cont | upperci_cont |
|---|---|---|---|---|---|---|---|---|---|---|---|---|
| PCE_CHD | 0.683 (0.676 - 0.69) | 164763 | 0.6204 | 0.6967 | 0.6943 | -0.014 | NA | NA | NA | NA | NA | NA |
| PCE_IRS_PGS003725 | 0.711 (0.704 - 0.717) | 164353 | 0.5877 | 0.7012 | 0.6977 | -0.014 | 0.0512 | 0.0044 | 0.0467 | 0.3228 | 0.0083 | 0.3145 |
| PCE_IRS_PGS003356 | 0.705 (0.698 - 0.711) | 164353 | 0.5718 | 0.7015 | 0.6974 | -0.014 | 0.0354 | 0.0047 | 0.0308 | 0.2496 | 0.0114 | 0.2382 |
| PCE_IRS_PGS004698 | 0.702 (0.696 - 0.709) | 164353 | 0.5710 | 0.6992 | 0.6952 | -0.015 | 0.0324 | 0.0024 | 0.0300 | 0.2646 | 0.0101 | 0.2545 |
| PCE_IRS_PGS004696 | 0.706 (0.7 - 0.713) | 164353 | 0.5815 | 0.6970 | 0.6933 | -0.014 | 0.0407 | 0.0002 | 0.0405 | 0.2890 | -0.0068 | 0.2958 |
| PCE_IRS_CHDMulti | 0.707 (0.7 - 0.713) | 164353 | 0.5796 | 0.7021 | 0.6982 | -0.014 | 0.0438 | 0.0053 | 0.0386 | 0.2781 | 0.0022 | 0.2759 |
| PCE_IS | 0.682 (0.675 - 0.689) | 164509 | 0.5396 | 0.6955 | 0.6903 | -0.015 | NA | NA | NA | NA | NA | NA |
| PCE_IRS_PGS002053 | 0.683 (0.676 - 0.689) | 164509 | 0.5374 | 0.6954 | 0.6901 | -0.015 | -0.0023 | -0.0001 | -0.0022 | 0.0323 | 0.0034 | 0.0289 |
| PCE_IRS_PGS004939 | 0.683 (0.677 - 0.69) | 164509 | 0.5439 | 0.6967 | 0.6916 | -0.014 | 0.0056 | 0.0012 | 0.0044 | 0.0478 | -0.0036 | 0.0515 |
| PCE_IRS_PGS000039 | 0.683 (0.676 - 0.689) | 164509 | 0.5417 | 0.6967 | 0.6915 | -0.014 | 0.0033 | 0.0011 | 0.0022 | 0.0319 | 0.0019 | 0.0300 |
| PCE_IRS_ISMulti | 0.687 (0.68 - 0.693) | 164509 | 0.5472 | 0.6974 | 0.6924 | -0.014 | 0.0096 | 0.0019 | 0.0076 | 0.1092 | -0.0051 | 0.1144 |
| PCE_AAA | 0.756 (0.744 - 0.768) | 164828 | 0.6953 | 0.6873 | 0.6874 | -0.03 | NA | NA | NA | NA | NA | NA |
| PCE_IRS_PGS002054 | 0.758 (0.746 - 0.77) | 164828 | 0.6970 | 0.6883 | 0.6884 | -0.029 | 0.0027 | 0.0010 | 0.0017 | 0.0534 | 0.0011 | 0.0524 |
| PCE_IRS_PGS001784 | 0.762 (0.75 - 0.774) | 164828 | 0.6953 | 0.6894 | 0.6895 | -0.029 | 0.0021 | 0.0021 | 0.0000 | 0.1109 | -0.0016 | 0.1124 |
| PCE_IRS_PGS003972 | 0.776 (0.764 - 0.788) | 164828 | 0.7185 | 0.6897 | 0.6899 | -0.029 | 0.0255 | 0.0024 | 0.0232 | 0.2962 | -0.0033 | 0.2996 |
| PCE_IRS_AAAMulti | 0.778 (0.766 - 0.79) | 164828 | 0.7270 | 0.6878 | 0.6881 | -0.029 | 0.0322 | 0.0005 | 0.0318 | 0.3410 | 0.0019 | 0.3391 |
| PCE_PAD | 0.727 (0.721 - 0.734) | 164763 | 0.6204 | 0.6967 | 0.6943 | -0.014 | NA | NA | NA | NA | NA | NA |
| PCE_IRS_PGS002055 | 0.728 (0.721 - 0.734) | 164763 | 0.6206 | 0.6972 | 0.6949 | -0.014 | 0.0007 | 0.0005 | 0.0002 | 0.0204 | 0.0009 | 0.0195 |
| PCE_IRS_PGS004940 | 0.731 (0.724 - 0.737) | 164763 | 0.6243 | 0.6976 | 0.6953 | -0.013 | 0.0048 | 0.0009 | 0.0039 | 0.1069 | 0.0120 | 0.0949 |
| PCE_IRS_PADMulti | 0.732 (0.726 - 0.739) | 164763 | 0.6233 | 0.7001 | 0.6977 | -0.013 | 0.0063 | 0.0034 | 0.0030 | 0.1214 | -0.0059 | 0.1273 |
| PCE_ASCVD | 0.702 (0.698 - 0.706) | 164150 | 0.5508 | 0.7171 | 0.7034 | 0.016 | NA | NA | NA | NA | NA | NA |
| PCE_IRS_ASCVDMulti | 0.705 (0.701 - 0.709) | 164150 | 0.5550 | 0.7179 | 0.7045 | 0.017 | 0.0049 | 0.0008 | 0.0042 | 0.1337 | 0.0005 | 0.1333 |
